# Place-based structural disadvantage and mortality inequality across 379 Polish counties, 2010–2025: a spatial Mundlak analysis

**DOI:** 10.64898/2026.11.02.26367044

**Authors:** Tomasz Madej, Bogdan Wojtyniak, Daniel Rabczenko, Anna Smaga, Stefan Boguslawski

## Abstract

Place-based mortality inequalities may persist even as short-term associations with local conditions change. We examined age-sex-standardised all-cause mortality in 379 harmonised Polish counties (2010–2025), asking whether durable between-county differences or lagged within-county deviations better characterise mortality geography. Spatial Mundlak generalised additive models allowed non-linear associations and county-specific spatial modification; a pre-COVID model (2010–2019) was compared with overlapping models through 2022 and 2025. In the pre-COVID model, median county average partial slopes per standard deviation were −26.1 deaths per 100,000 for higher education, +29.4 for primary/lower-secondary education and +30.2 for PM2.5. Directions of these between-county gradients persisted across analytical windows. By contrast, within-county associations were less stable and sometimes reversed sign in pandemic-inclusive models. County-level uncertainty indicated stronger and more coherent pre-COVID spatial patterns for education, PM2.5 and population density than for several health-care and migration terms. Strict leave-one-year-out validation revealed weak temporal transportability, particularly for the 2010–2022 model, limiting predictive interpretation. Outcome-defined comparisons further showed that educational composition remained an important separator of high- and low-mortality counties after COVID-19, while the fitted PM2.5 contribution contrast diminished. These findings distinguish durable configurations of place-based disadvantage from period-dependent ecological co-movement. They support geographically sensitive, multi-domain approaches to reducing mortality inequalities, while not establishing individual-level causal effects.

## 1. Introduction

Health inequalities are socially and geographically patterned through places as well as through individual circumstances. Local labour markets, educational infrastructure, health-care capacity, environmental quality, housing and institutional legacies cluster geographically, creating configurations of advantage and disadvantage that are closely linked to population health across generations (Macintyre et al., 2002; Cummins et al., 2007; Diez Roux and Mair, 2010). Place-based approaches have proliferated in part because the spatial clustering of social determinants makes it impossible to separate individual exposures from the area contexts in which they are embedded and through which they operate.

Poland offers an informative setting. The country achieved substantial national mortality improvements after 1989, but these gains have been distributed unequally across its 379 counties (powiaty), which differ markedly in educational composition, employment, health-care capacity, urbanisation, migration and PM2.5 air pollution (Wojtyniak and Gorynski, 2020, 2022). This geographic heterogeneity reflects post-socialist economic restructuring, uneven regional development and historical legacies spanning nineteenth-century partitions, post-war settlement and the rapid dissolution of state-owned agriculture after 1991 (Bąk et al., 2021; Grosfeld and Zhuravskaya, 2015). These histories provide interpretive context but are not encoded as exposures in our models. COVID-19 then created a distinct temporal break in mortality trends, mobility, health-care use and labour-market conditions.

Two analytical challenges motivate our approach. First, durable differences between places are not equivalent to short-run changes within the same place. A county characterised by chronically low educational attainment occupies a different structural position than one that temporarily falls below its own historical level. Pooled panel coefficients blend these sources of variation; county fixed effects eliminate the persistent between-county gradient that is central to place-based inequality research. The Mundlak correlated-random-effects decomposition retains both components and estimates them simultaneously (Mundlak, 1978; Bell and Jones, 2015; Arkhangelsky and Imbens, 2024). This separation matters because annual county indicators can co-move with unobserved changes in behaviour, composition and service use; allowing short-run ecological co-movement to be estimated separately rather than conflated with the structural place gradient.

Second, the mortality association of a given social or environmental determinant need not be spatially uniform. Uniform-coefficient models impose a national average relationship even when institutional, environmental and social conditions differ substantially across space. Spatially varying-coefficient approaches can reveal contextual heterogeneity in exposure–health relationships (Janko et al., 2019; Comber et al., 2023, 2024a, 2024b), and recent guidance highlights the importance of explicit specification checks, perturbation sensitivity and local uncertainty quantification (Comber et al., 2026; Irekponor and Oshan, 2025).

This study treats place not simply as the location of exposure but as a durable configuration of social, environmental and service conditions. Poland provides a setting in which post-socialist restructuring, territorial unevenness and the COVID-19 disruption can be studied together within harmonised local administrative units. Our substantive question is whether the geography of mortality inequality remains anchored in persistent differences between counties even when within-county associations change during a major societal shock. We examine this question using spatial Mundlak generalised additive models (GAMs) for 379 counties over 2010–2025, guided by the Dahlgren–Whitehead framework as a heuristic rather than a causal diagram. We ask: (1) how durable place differences compare with lagged local deviations; (2) where education, employment, service-capacity and pollution gradients vary geographically; (3) whether the principal place gradients persist when COVID-19 and post-COVID years are included; and (4) which fitted contributions distinguish counties with high versus low observed mortality. The broader contribution is to show why stable spatial inequality and unstable temporal association can coexist, and what that distinction means for interpreting place-sensitive health policy.

## 2. Data and measures

### 2.1 Study area and analytical panel

The unit of analysis is the Polish county (powiat), the second-order administrative unit below NUTS-3 subregions. Counties approximate local labour-market and service environments while remaining small enough to reveal sub-regional variation. Wałbrzych regained city-with-county-rights status on 1 January 2013, increasing the contemporary number of county-level units to 380. To maintain a spatially consistent panel across 2010–2025, Wałbrzych city and the surrounding Wałbrzych county were harmonised to a common pre-change geography, yielding 379 analytical units throughout. The primary pre-COVID model covers 2010–2019; two overlapping extensions cover 2010–2022 and 2010–2025, adding COVID-period and post-COVID controls. Because within-county determinants are lagged by one year, effective estimation samples begin in 2011: n=3,411 (Model 1), n=4,548 (Model 2), and n=5,685 (Model 3).

### 2.2 Mortality outcome

The outcome was the annual age-sex standardised all-cause mortality rate (ASMR) per 100,000 residents. County deaths and population denominators were obtained from Statistics Poland’s Local Data Bank (Statistics Poland, 2026). Deaths were harmonised into 18 age groups (0–4 through 85+) separately for women and men. Direct standardisation used a fixed sex-by-age reference distribution pooled across counties in 2010–2019 and applied uniformly from 2010 to 2025. Further construction and harmonisation details are provided in Supplementary Methods S1.2 and Supplementary Table S2.

### 2.3 Determinants

Nine between-county and eight lagged within-county determinants were derived from Statistics Poland’s Local Data Bank, the 2011 and 2021 national population censuses, and CAMS air-quality reanalysis products (Supplementary Table S2; raw determinant geography and annual change: Supplementary Figures S6–S7). Educational composition (higher education share; primary/lower-secondary education share) was constructed by linear interpolation between census anchors and should therefore be interpreted as a structural trajectory rather than an independently observed annual shock. The two education indicators represent the upper and lower tails of the educational distribution rather than complementary categories; upper-secondary, vocational and post-secondary non-tertiary education form a substantial intermediate group. Annual mean PM2.5 was derived from CAMS EAC4 (2010–2012) and the CAMS European air-quality reanalysis (2013 onward) (Copernicus Atmosphere Monitoring Service, 2026a, 2026b). Distance to the nearest city with county rights entered as a time-invariant structural marker. All time-varying determinants were standardised to a common 2010–2019 reference scale. Supplementary Table S1 maps each determinant to its Dahlgren–Whitehead layer, Mundlak role and interpretive caveat. Additional details on the census-derived education series and PM2.5 construction are provided in Supplementary Methods S1.3–S1.4.

## 3. Analytical strategy

### 3.1 Mundlak decomposition

For each time-varying determinant, the lagged county-year value was decomposed into a persistent county mean and a lagged within-county deviation: where is the determinant observed for county in the preceding year, is the county-specific mean of that determinant over the relevant analytical window, and is the lagged within-county deviation from that mean.

The county mean represents persistent between-county differences; the lagged deviation represents a year-specific departure from the county’s own typical level. Distance to a city with county rights entered only as a between-county term. The decomposition therefore produces nine between-county and eight within-county components.

A pooled model blends persistent spatial inequality with short-run ecological co-movement; county fixed effects discard the stable place differences central to this analysis. Mundlak retains both and makes their divergence an empirical result. We interpret within-county terms as historically contingent ecological associations that can reflect unmeasured changes in behaviour, composition, care-seeking or local service provision. Supplementary Methods S1.9 formalises this interpretive hierarchy. The conceptual distinction between structural and within-county layers is summarised in Supplementary Methods S1.1, with the interpretive hierarchy for within terms in S1.9.

### 3.2 Spatial Mundlak GAM

The outcome was modelled using a Gaussian identity-link GAM:

Here is the smooth calendar-time term, contains COVID-period indicators, is the baseline spatial mortality surface, is the national non-linear partial function for Mundlak component, and is its spatially varying coefficient modifier. Determinant-specific national functions and spatial modifiers used shrinkage thin-plate splines; the temporal and baseline spatial terms used thin-plate regression splines. All analyses were performed in R and estimated with mgcv’s bam using fREML and (Wood, 2017).

### 3.3 Basis-dimension selection

Basis dimensions were selected empirically using a 48-model grid on the pre-COVID model, spanning for one-dimensional determinant smooths, for SVC surfaces, for the baseline spatial surface, and for calendar time. Selection targeted the smallest dimensions for which curves, average partial slopes and SVC surfaces stabilised under upward increases in ; an additional 175→200 boundary check confirmed the stability of for the baseline surface. Final dimensions (global:, SVC:, baseline:, year:) were fixed across Models 1–3 to preserve cross-window comparability (Supplementary Methods S1.5; Supplementary Table S3).

### 3.4 Effect summaries and local uncertainty

For each Mundlak component, effect curves varied the focal component across 25 values spanning its empirical P2–P98 range while holding all non-focal determinant components at zero. For the focal determinant, both its national smooth and its corresponding spatially varying coefficient term were retained. Average partial slopes (APS) are finite-difference derivatives averaged over the grid within each county and summarised by the median and IQR across counties; IQRs describe fitted spatial heterogeneity, not sampling uncertainty. Marginal-effect maps show the local slope per +1 SD. Between-county accumulated-contribution maps compare each county’s structural level with the 0-SD reference; within-county maps compare a +1-SD lagged deviation with 0. These quantities answer different questions and can have opposite signs under a non-linear response surface (Supplementary Methods S1.11). Local uncertainty was propagated from the GAM coefficient covariance matrix using approximate pointwise 95% intervals; counties were classified as robust positive, robust negative or uncertain (Mennis, 2006; Irekponor and Oshan, 2025; Supplementary Methods S1.10; Supplementary Table S9 and Figures S11–S12).

To quantify the role of spatial modification, we performed a same-fit post-estimation sensitivity in which the determinant-specific spatial modifier for each focal component was set to zero while all other fitted model components were retained. This contrasts the reported focal relationship (national smooth plus spatial modifier) with the national smooth alone; no alternative model was refitted. Results for all three analytical windows are reported in Supplementary Table S10 and Supplementary Methods S1.13.

### 3.5 Linear benchmark and validation

A linear Mundlak ridge model with the same decomposition served as a homogeneous benchmark; uncertainty was estimated using 1,000 recentred dependent-multiplier bootstrap replications (Cameron et al., 2008; Shao, 2010; Zhang and Politis, 2022). Validation combined basis-dimension checks, annual Moran’s I, county-level lag-1 residual autocorrelation, and strict leave-one-year-out cross-validation with preprocessing recomputed within each training fold. A structured-noise perturbation stress test (50 replicates per model) summarised stability using curve-shape correlations, APS sign agreement, signal-to-noise ratios and, where meaningful, spatial-pattern correlations (Supplementary Methods S1.6–S1.7; Supplementary Tables S4–S6).

### 3.6 Post-estimation profile analysis

To relate fitted determinant contributions to substantively recognisable county profiles, we used the common Model 3 fit but defined comparison groups independently of the model. Within each descriptive period (2010–2019, 2020–2022 and 2023–2025), counties in the bottom and top deciles of observed mean ASMR were classified as LOW and HIGH, respectively. We first compared raw county characteristics (Supplementary Table S7) and then determinant-specific accumulated contributions from Model 3 (Supplementary Table S8). HIGH-minus-LOW contrasts are differences in reference-based fitted contributions, not marginal slopes or causal attributable effects. A separate sensitivity analysis defines groups by total model-derived determinant burden (Supplementary Methods S1.8; Supplementary Figure S10).

## 4. Results

### 4.1 Mortality geography, model fit and temporal transportability

Median county ASMR was 1,060.9 per 100,000 (IQR 987.5–1,110.3) in 2010–2019, rose to 1,193.7 (IQR 1,114.0–1,250.6) during 2020–2022, and fell to 961.7 (IQR 898.5–1,016.5) in 2023–2025—below the pre-pandemic median (Figure 1). The pandemic increase was geographically widespread; the post-pandemic decline was spatially uneven.

**Figure 1.**
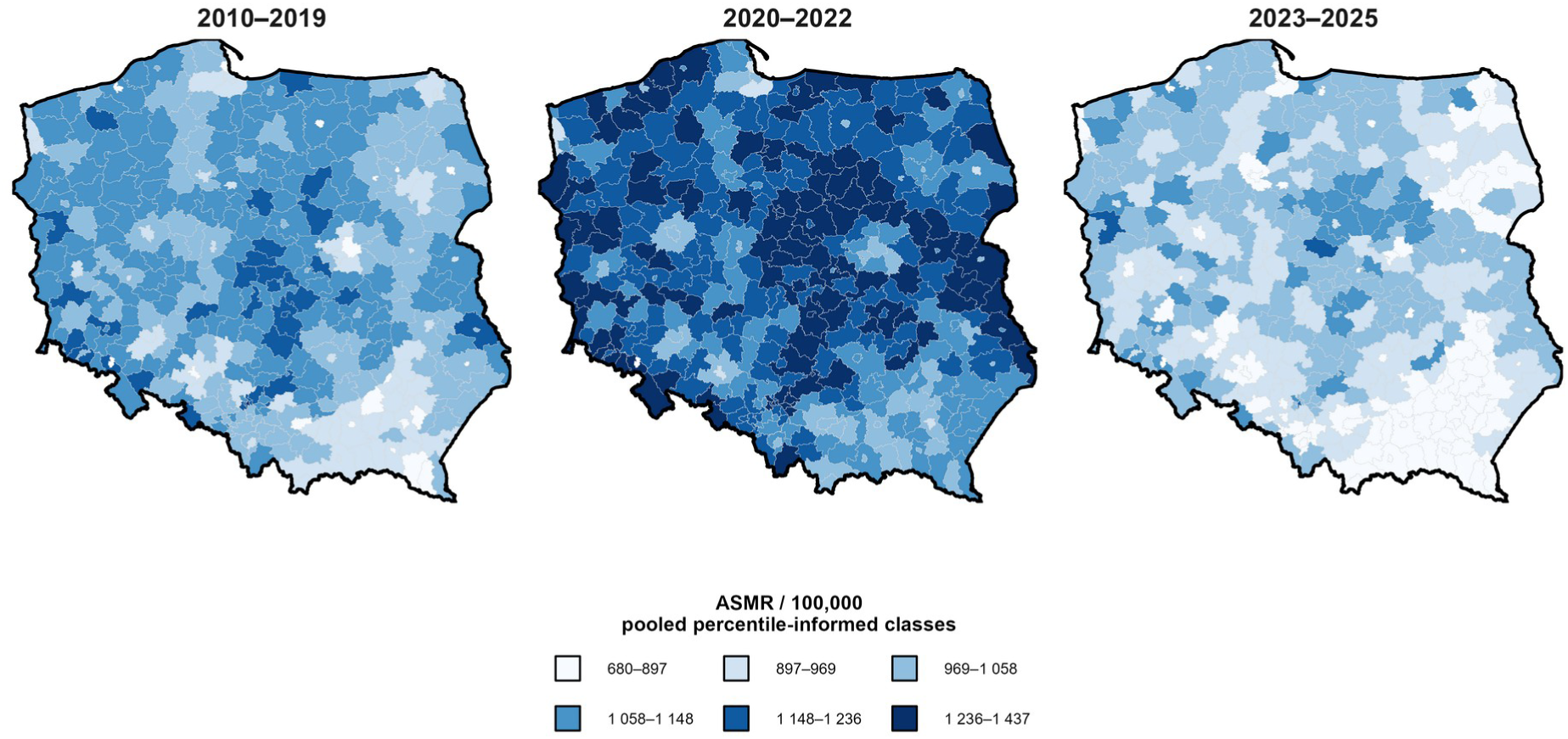
Descriptive geography of age-sex standardised mortality across three periods. Maps show county mean ASMR per 100,000 for 2010–2019, 2020–2022 and 2023–2025 using one pooled percentile-informed class scheme so colours are comparable across panels. Map lines delineate study areas and do not necessarily depict accepted national boundaries.

The spatial Mundlak GAM explained 73.1% of variance in Model 1 (deviance explained 74.4%; RMSE 49.8 per 100,000; Table 1). In-sample RZ was 0.803 in Model 2 and 0.787 in Model 3. Positive residual Moran’s I occurred in 0/9 model years in Model 1, 1/12 in Model 2 and 4/15 in Model 3 (full diagnostics: Supplementary Table S4). Strict leave-one-year-out validation showed marked temporal instability after 2020: median held-out RZ was 0.479 for Model 1, −3.544 for Model 2 and +0.326 for Model 3, while mean held-out RMSE remained large in both pandemic-inclusive windows. We therefore treat the GAMs as period-conditioned explanatory models rather than forecasting tools.

**Table 1.**
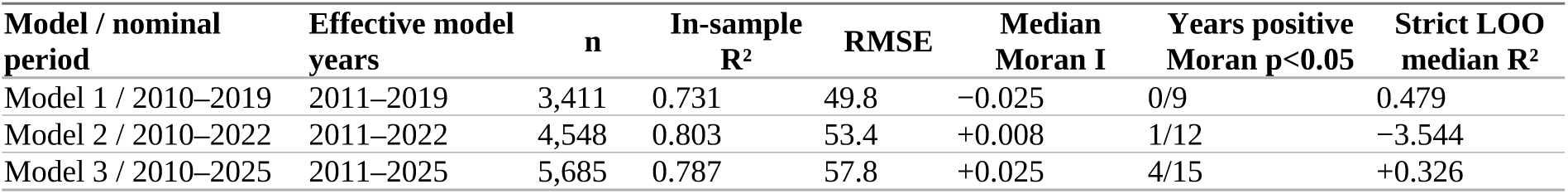
Spatial Mundlak GAM fit and validation across the three overlapping analysis periods. RMSE is in ASMR per 100,000. Moran counts refer to annual one-sided positive residual spatial autocorrelation at p<0.05. Strict LOO = leave-one-year-out cross-validation with preprocessing recomputed within the training fold.

### 4.2 Persistent between-county structural gradients

The largest pre-COVID associations were concentrated in the between-county layer (Figure 2; Table 2). In Model 1, PM2.5 had an APS of +30.2 ASMR per 100,000 per SD (county IQR +16.3 to +43.6), primary/lower-secondary education +29.4 (+16.1 to +37.9), and higher education −26.1 (−46.2 to −5.7). Population density was +15.1 and long-term unemployment +7.5. Gross wages had a small negative median slope (−2.6); physicians, net migration and distance to city were close to zero.

**Figure 2.**
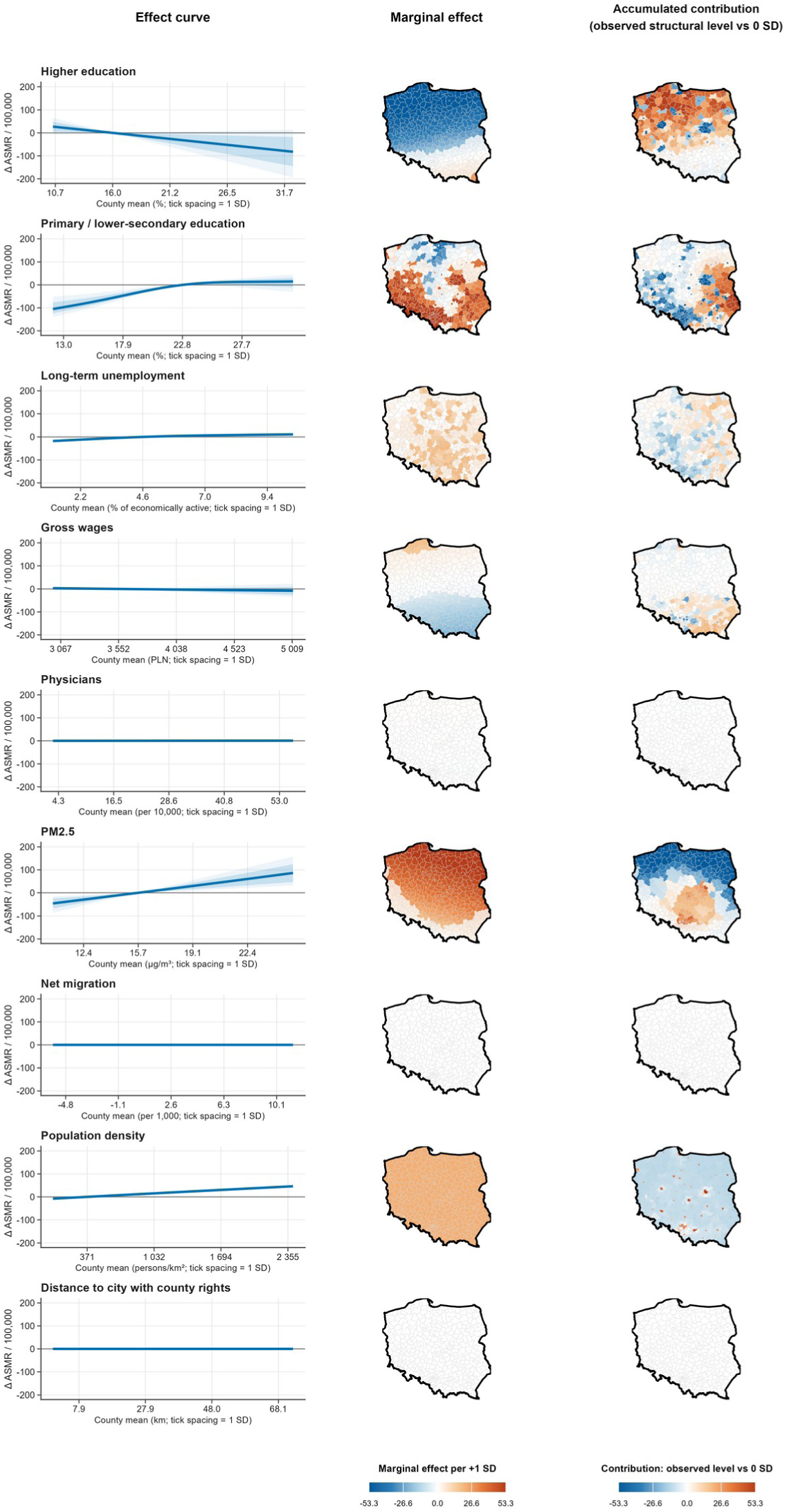
Between-county structural effects in the primary pre-COVID spatial Mundlak GAM (2010–2019; effective model years 2011–2019). Left: county-specific partial effect curves summarised by median, IQR and P10–P90 bands; the bands show fitted cross-county heterogeneity, not confidence intervals. Centre: local marginal effects per +1 SD at each county’s observed structural level. Right: accumulated contributions of observed structural levels relative to the 0-SD reference. Pale counties indicate approximate pointwise 95% intervals that include zero. Colour limits are symmetric and common across paired cartograms, with robust pooled clipping used only for visual scaling. uncertainty maps: Supplementary Figure S11. Quantities are fitted partial contrasts, not causal effects. Map lines delineate study areas and do not necessarily depict accepted national boundaries.

**Table 2.** GAM average partial slopes across analysis periods, by Mundlak layer. Values are median county average partial slopes [county IQR], expressed as ΔASMR per 100,000 for a +1-SD change on the common Model-1 reference scale. IQRs describe spatial heterogeneity, not sampling-confidence intervals. ≈₀ denotes effects below ±0.05 per SD.

| **Determinant** | **Layer** | **Model 1 2010–2019** | **Model 2 2010–2022** | **Model 3 2010–2025** |
|---|---|---|---|---|
| Higher education | Between | −26.1 [−46.2, −5.7] | −25.0 [−43.2, −7.0] | −33.5 [−46.2, −20.7] |
| Primary/lower-secondary education | Between | +29.4 [+16.1, +37.9] | +24.2 [+14.2, +35.6] | +19.2 [+7.5, +33.3] |
| Long-term unemployment | Between | +7.5 [+6.3, +8.9] | +8.9 [+5.7, +11.1] | +9.6 [+9.3, +9.9] |
| Gross wages | Between | −2.6 [−8.6, +3.0] | −1.4 [−6.1, +3.9] | −3.5 [−7.0, −0.4] |
| Physicians | Between | +0.2 [−0.3, +0.7] | +2.8 [−3.5, +9.1] | +9.9 [+2.3, +18.4] |
| PM2.5 | Between | +30.2 [+16.3, +43.6] | +31.6 [+19.0, +44.0] | +36.7 [+14.3, +61.3] |
| Net migration | Between | ≈0 | ≈0 | ≈0 |
| Population density | Between | +15.1 [+15.1, +15.1] | +14.1 [+14.1, +14.1] | +2.5 [−0.4, +5.1] |
| Distance to city | Between | ≈0 | ≈0 | +0.7 [+0.7, +0.7] |
| Higher education | Within | ≈0 | −8.7 [−8.7, −8.7] | +2.0 [−0.5, +3.7] |
| Primary/lower-secondary education | Within | +7.1 [+3.4, +10.7] | +4.4 [−0.9, +8.9] | +7.0 [+2.0, +12.2] |
| Long-term unemployment | Within | −1.1 [−1.1, −1.1] | −5.2 [−5.9, −4.1] | −5.5 [−8.2, −3.1] |
| Gross wages | Within | ≈0 | −0.5 [−2.2, +1.4] | −1.1 [−2.8, +1.0] |
| Physicians | Within | ≈0 | +0.3 [−0.1, +0.7] | ≈0 |
| PM2.5 | Within | ≈0 | +4.6 [+0.8, +8.2] | −13.2 [−16.0, −9.3] |
| Net migration | Within | ≈0 | +0.5 [−0.1, +1.1] | +1.3 [+0.2, +2.2] |
| Population density | Within | +3.7 [+3.7, +3.7] | +0.6 [+0.6, +0.6] | +0.8 [+0.8, +0.8] |

These gradients persisted as the analytical window expanded. Higher education remained negative (−25.0 in Model 2; −33.5 in Model 3), while primary/lower-secondary education (+24.2; +19.2), long-term unemployment (+8.9; +9.6) and PM2.5 (+31.6; +36.7) remained positive. Population density weakened from +15.1 in Model 1 to +2.5 in Model 3, whereas the physician APS increased from near zero to +9.9. Cross-period APS are shown in Supplementary Figure S1; complete between-county effect grids for Models 1–3 are in main-text Figure 2 and Supplementary Figures S2 and S4.

Local uncertainty materially qualified the pre-COVID spatial maps. Approximate pointwise intervals supported a negative higher-education marginal effect in 65.7% of counties, a positive primary/lower-secondary education effect in 55.4%, a positive PM2.5 effect in 72.3%, and a positive population-density effect in all counties; physicians, net migration and distance to city had no counties with a robust local marginal sign. Selected coverage percentages are reported in Supplementary Table S9 and full Model 1 uncertainty maps in Supplementary Figures S11–S12. These intervals are pointwise and model-conditional, not multiplicity-adjusted spatial significance tests.

The linear benchmark agreed with the GAM on the broad direction of education, unemployment, wages and PM2.5 but disagreed for population density and physicians (Figure 3; full ridge results: Supplementary Table S5)—expected when a homogeneous coefficient averages across exposure ranges and locations for which the GAM can estimate different relationships.

**Figure 3.**
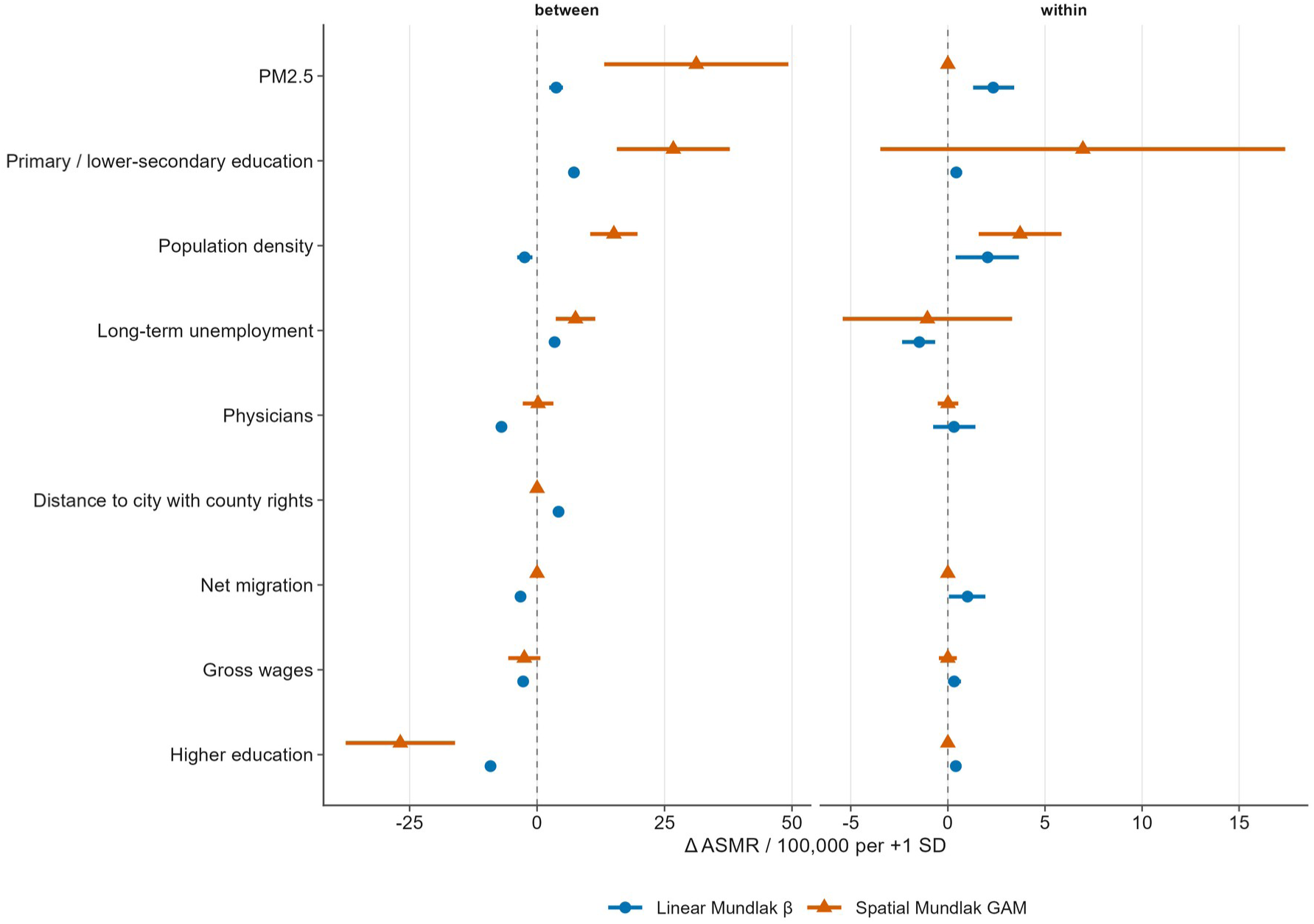
Linear Mundlak benchmark and spatial GAM partial slopes, primary pre-COVID model (2010–2019). Blue circles show debiased linear Mundlak coefficients with 95% dependent-multiplier bootstrap confidence intervals (B=1,000). Orange triangles show the GAM mean average derivative with approximate 95% coefficient-covariance intervals derived from the fitted GAM. These intervals quantify estimation uncertainty and should not be interpreted as the cross-county heterogeneity reported by the APS IQRs in Table 2. Discrepancies can reflect non-linearity, spatially varying slopes and differential allocation of shared spatial signal. Full ridge estimates are in Supplementary Table S3.

### 4.3 Within-county lagged dynamics and ecological co-movement

Within-county lagged associations were generally smaller than between-county gradients and changed more across estimation windows (Figure 4; Table 2). In the pre-COVID model, primary/lower-secondary education (+7.1) and population density (+3.7) had the clearest positive APS values; long-term unemployment was modestly negative (−1.1); remaining within terms were close to zero.

**Figure 4.**
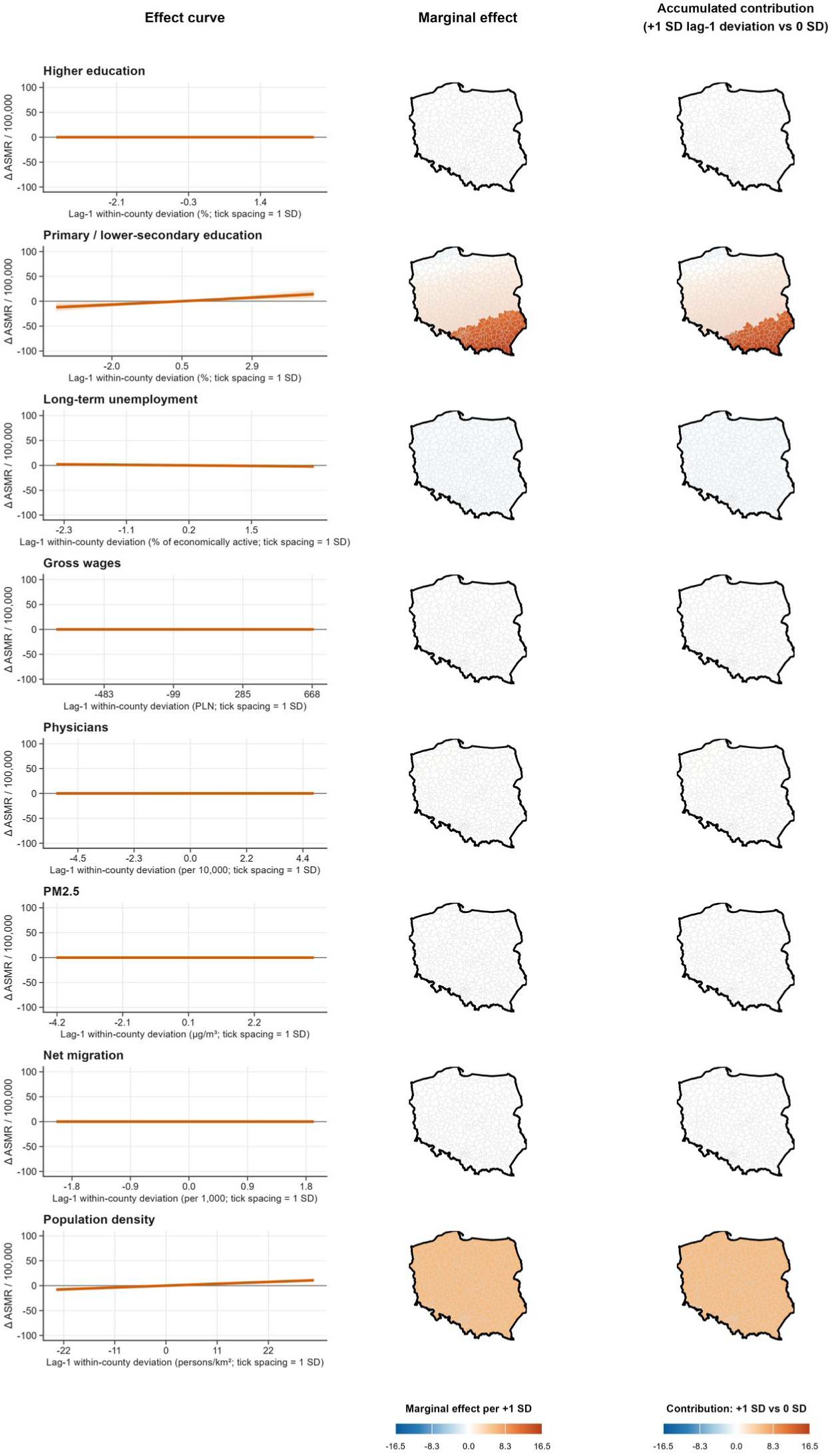
Within-county lag-1 effects in the primary pre-COVID spatial Mundlak GAM (2010–2019). Layout follows Figure 2. Within effects describe conditional temporal coupling relative to each county’s own historical level and may track unmeasured behavioural, compositional or service changes. Education within terms use census-derived annual trajectories. Colour limits are symmetric and common across paired cartograms, with robust pooled clipping used only for visual scaling. Map lines delineate study areas and do not necessarily depict accepted national boundaries.

Pandemic-inclusive models produced qualitatively different within-county patterns. Higher education was negative in Model 2 (−8.7) but close to zero/weakly positive in Model 3 (+2.0); PM2.5 was positive in Model 2 (+4.6) but negative in Model 3 (−13.2); long-term unemployment was negative in both (−5.2 and −5.5). These reversals should not be interpreted as established risk factors becoming protective. They indicate that short-term county deviations were embedded in a changed temporal regime in which employment, mobility, energy use, health-care utilisation and mortality co-moved differently. Complete within-county effect grids for Models 1–3 are in main-text Figure 4 and Supplementary Figures S3 and S5; Supplementary Methods S1.9 and S1.14 provide the interpretation hierarchy for such sign changes.

Structured-noise perturbation supported the stability of the major between-county curves more consistently than several local spatial modifiers and within-county surfaces. Education and PM2.5 between-county curves showed high signal-to-noise ratios and near-unit shape correlations, whereas some within-county and spatial-pattern metrics were more variable (Supplementary Table S6).

### 4.4 What distinguishes high- and low-mortality counties?

In 2010–2019, median ASMR was 1,168.7 per 100,000 in the highest observed-ASMR decile versus 901.4 in the lowest. High-mortality counties had less higher education (14.1% versus 24.3%), more primary/lower-secondary education (23.9% versus 14.5%), fewer physicians (13.4 versus 35.7 per 10,000), much lower population density (80 versus 1,091 persons/kmZ) and greater distance from a city with county rights (27.7 versus 0.0 km). The same outcome-defined profiling is reported for the COVID and post-COVID periods in Supplementary Table S7.

Model 3 contribution profiles show how the fitted structural configuration separating HIGH and LOW counties changed across descriptive periods. In the pre-COVID window, HIGH-minus-LOW mean contribution contrasts were +68.9 ASMR per 100,000 for higher education, +58.0 for primary/lower-secondary education, +22.1 for PM2.5, +5.3 for long-term unemployment and +4.0 for wages. Education remained the clearest separator during COVID (+79.3 and +52.0) and post-COVID (+58.5 and +48.6), whereas the PM2.5 contrast attenuated to +8.2 during COVID and +0.3 post-COVID (Supplementary Table S8). Figure 5 shows the Model 1 dominant-contribution map; Supplementary Figures S8–S9 show Models 2–3. Dominance is relative: the map identifies the largest accumulated contribution within a county, not the determinant with the largest national slope. Consequently, a factor such as distance to a city can become visually dominant in Model 3 even though its national APS is small (+0.7) and its spatial modifier is effectively shrunk to zero; this reflects the reference-based contrast and smaller competing contributions rather than the emergence of a strong post-COVID distance effect. Supplementary Figure S10 provides the model-burden grouping sensitivity.

**Figure 5.**
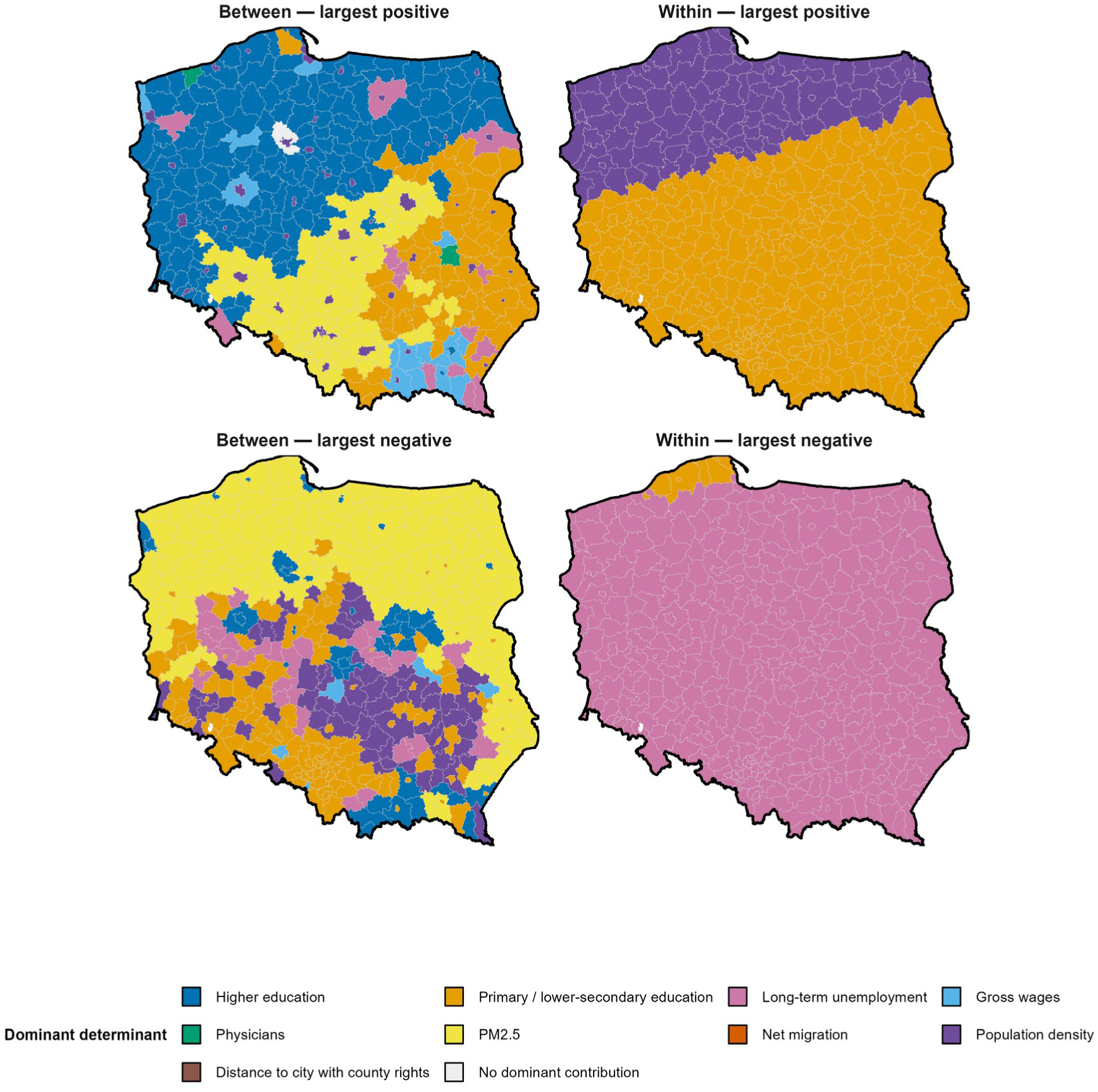
Dominant accumulated determinant contribution by county in the primary pre-COVID spatial Mundlak GAM (2010–2019). Top row: determinant with the largest positive accumulated contribution; bottom row: determinant with the largest negative accumulated contribution. Left column: between-county structural layer; right column: within-county lag-1 layer. Grey indicates no dominant contribution. Dominance identifies the largest reference-based accumulated contribution within each county and does not rank determinants by national effect magnitude. A determinant with a small average partial slope may therefore appear as locally dominant when its observed level lies far from the 0-SD reference or when competing determinant contributions are smaller. Accumulated-contribution signs need not match local marginal slopes under non-linearity. Map lines delineate study areas and do not necessarily depict accepted national boundaries.

## 5. Discussion

### 5.1 Structural mortality inequality as a layered property of place

The most robust finding is that persistent between-county structural differences—in education, PM2.5 and labour-market conditions—were larger and more temporally stable than most lagged within-county associations. This pattern is consistent with a layered determinants perspective in which disadvantage is bundled across educational, labour-market, environmental, service and accessibility domains rather than attributable to a single exposure (Dahlgren and Whitehead, 1991, 2021). The stability of the leading between-county ordering despite pandemic-era disruption is therefore more informative than any individual short-run sign reversal.

Education should not be read as an isolated schooling effect. County educational composition indexes material resources, labour-market opportunities, health literacy, health-related behaviours and capacity to navigate services. Polish and European evidence links educational position to health behaviour, preventive orientation, health literacy and mortality (Stelmach et al., 2004; Puchalski and Korzeniowska, 2011; Huijts et al., 2017; Laaksonen et al., 2008; Stormacq et al., 2019; IHME-CHAIN Collaborators, 2024). The non-linear curves and spatially varying slopes further imply that a common 1-SD contrast does not carry an identical association across the exposure range or across counties.

The two education indicators capture different parts of the county educational distribution. Higher-education share may index concentrations of human capital, professional employment, institutional resources, health literacy, social networks and health-related norms; area-level college-education composition has been associated with mortality beyond individual attainment in other settings (Bor et al., 2024). By contrast, a high primary/lower-secondary share marks the lower tail of the distribution and is better interpreted as accumulated structural disadvantage, labour-market exclusion and limited socioeconomic mobility. European evidence also indicates that educational expansion can make the remaining low-education group increasingly selected, changing the magnitude of education–mortality inequalities (Mackenbach et al., 2015; Östergren et al., 2017). Upper-secondary, vocational and post-secondary non-tertiary education form a substantial intermediate category, so the two shares are not mirror images.

PM2.5 is a measured environmental exposure rather than a generic deprivation proxy. Its large positive between-county APS persisted across all analytical windows and its southern concentration is consistent with the documented particulate-pollution burden of Upper Silesia and adjacent southern Poland (Kobza et al., 2018). By contrast, the within-PM2.5 association changed sign across pandemic-inclusive windows (+4.6 in Model 2; −13.2 in Model 3). That reversal is not credible evidence of a protective pollution effect. It is more plausibly a marker of altered temporal coupling during a period when mortality shocks, mobility, heating and emissions changed simultaneously.

The apparently weak conditional slopes for some service and accessibility indicators require similar care. Physicians, migration and distance to a city are strongly patterned components of county population geography, but much of their variation overlaps with education, density, labour markets and urbanity. A small average partial slope therefore does not imply substantive irrelevance. This distinction is especially visible in the dominant-contribution maps: distance to a city appears frequently as a Model 3 dominant accumulated contribution even though its average slope remains small and its determinant-specific spatial modifier is effectively penalised to zero. Dominance here is relative to other reference-based contributions, not evidence that distance suddenly became the strongest post-COVID mortality determinant.

County wage levels likewise index local labour-market composition and productivity rather than household disposable income, while net migration may capture selective demographic sorting and the relative attractiveness of employment, housing and service environments. The indicators should therefore be interpreted as partially overlapping markers of place-based domains. Their conditional coefficients describe what remains after the other modelled dimensions of place are held constant; they are not isolated causal effects.

### 5.2 Within-county dynamics: temporal contingency, not noise

The within-county results warrant interpretation rather than dismissal, but their meaning differs fundamentally from the between-county layer. A within term asks whether a county’s departure from its own typical determinant level co-varies with next-year mortality after conditioning on persistent structure and the other model components. The pandemic altered this temporal regime. Several within terms changed sign or magnitude while the leading structural ordering persisted, indicating historically contingent ecological co-movement rather than a reversal of the underlying long-run determinants.

Strict leave-one-year-out results reinforce this interpretation. Model 2 showed extremely poor temporal transportability, while Model 3’s median held-out RZ recovered to a positive value but prediction errors remained large. These models are therefore period-conditioned explanatory surfaces, not forecasting models, and differences across the overlapping windows should not be read as clean causal pre/post contrasts.

The post-COVID period nevertheless suggests a partial reweighting of the structural profile associated with high mortality. Determinants that strongly differentiated HIGH from LOW counties before the pandemic did not contribute equally strongly afterwards: education remained the dominant separator, whereas the PM2.5 HIGH-minus-LOW contribution contrast fell from +22.1 before COVID to +0.3 in 2023–2025. This does not mean that the underlying PM2.5 gradient disappeared—the between-county APS remained large—but that PM2.5 no longer separated the contemporaneous mortality extremes in the same way. Several mechanisms could contribute to the instability of within-county signs more generally: pandemic-era co-movement of employment, mobility, energy use and care utilisation; selective mortality or mortality displacement among frailer populations; secular improvement and compressed variability in exposures such as PM2.5; and a lower signal-to-noise ratio when remaining within-country variation is small relative to measurement error, shared trends and residual confounding. These mechanisms are not identified directly by the present ecological models and should be treated as hypotheses, not explanations established by the data (Supplementary Methods S1.14). In this sense, the post-COVID profile reflects a reconfiguration rather than a disappearance of structural mortality disadvantage.

### 5.3 Methodological contribution and policy implications

For place-based health research, the value of the spatial Mundlak approach lies in keeping persistent contextual configurations distinct from short-run ecological co-movement, while allowing associations to differ in shape and location. The method does not turn county-level associations into individual-level causal effects. National curves, local modifiers and model-based uncertainty jointly indicate which geographic patterns merit substantive interpretation and which remain weakly identified.

The policy implication is not that counties should be ranked by individual coefficients. Rather, high-mortality counties combine disadvantages across education, labour markets, health-care capacity and peripherality, while the relative contribution of specific domains can change over time. This supports multi-domain place-sensitive responses: educational and vocational investment, equitable primary and specialist health-care access, transport connectivity and continued air-quality improvement. The persistence of educational contrasts after COVID and the attenuation of PM2.5 as a HIGH–LOW separator also suggest that intervention priorities should be informed by both durable structural gradients and the changing composition of local mortality disadvantage.

The spatial pattern should also be read against Poland’s historically uneven regional development. Contemporary county differences overlay territories shaped by different nineteenth-century institutional and infrastructural regimes, post-war population transfers and industrialisation, the concentration and later dissolution of state agriculture in parts of northern and western Poland, and highly uneven post-1989 restructuring (Bąk et al., 2021; Grosfeld and Zhuravskaya, 2015; Herbst, 2023). These histories are not modelled exposures and the present surfaces cannot attribute mortality gradients to former partitions or later restructuring. They provide an interpretive backdrop for why education, labour-market conditions, migration, urban accessibility and service capacity remain geographically bundled (Supplementary Methods S1.12).

### 5.4 Limitations

This analysis is ecological and cannot identify individual-level effects. The Mundlak decomposition separates between- and within-county variation but does not eliminate time-varying confounding or unmeasured stable heterogeneity. Health behaviours, psychosocial stress, detailed service use and occupational exposures are incompletely observed; county indicators may partly capture unmeasured intermediary or compositional processes. Education values are census-derived trajectories rather than independently observed annual shocks.

PM2.5 changes source product in 2013; harmonisation reduces but cannot eliminate this discontinuity. The post-COVID descriptive window contains only three years (2023–2025), limiting inference about whether observed reweighting will persist. The three model windows overlap, so changes in APS across Models 1–3 are sensitivity comparisons rather than isolated pre-, COVID- and post-COVID effects. Selective mortality, mortality displacement and compressed exposure variability are plausible interpretations of some short-run sign changes but are not directly tested. Spatial concurvity is structurally expected; penalisation, basis sensitivity analysis, perturbation and coefficient-covariance uncertainty propagation reduce over-reading of unstable surfaces but do not make smooth components causally separable. Estimates remain regularised, model-conditional ecological associations.

## 6. Conclusion

Across 379 harmonised Polish counties, persistent educational and environmental differences between places were more stable correlates of mortality inequality than lagged within-county deviations over 2010–2025. This distinction matters beyond Poland: a major mortality shock can disrupt short-run ecological associations without erasing the underlying geography of structural disadvantage. Place-sensitive policy should therefore consider bundles of educational opportunity, labour-market conditions, health-care access and environmental quality rather than rank counties by isolated model coefficients. The estimates remain observational, ecological and model-conditional; they identify geographic patterns for further investigation, not causal effects of individual determinants.

## Data Availability

The harmonised analytical county-year panel and its documentation have been prepared for deposit in Zenodo, and the analysis code is maintained in a version-controlled repository. Public access and persistent identifiers are planned after acceptance, subject to verification of the redistribution terms of third-party source data.

## Ethics statement

This study used publicly available aggregate county-level administrative, demographic and environmental data. No identifiable individual-level information was used and no human participants were directly involved. Ethical approval and informed consent were not required.

## Data and code availability

County-level mortality and population data were obtained from Statistics Poland’s Local Data Bank; PM2.5 was derived from Copernicus Atmosphere Monitoring Service reanalysis products. Detailed data provenance and processing methods are reported in Supplementary Table S2. The harmonised analytical county-year panel and its documentation have been prepared for deposit in Zenodo, and the analysis code is maintained in a version-controlled repository. During double-anonymised review, author-identifying repository links and unpublished repository identifiers are withheld from the manuscript. Neither the dataset nor the code is currently available through a public archival DOI. Public access and persistent identifiers are planned after acceptance, subject to verification of the redistribution terms of third-party source data.

## Declaration of generative AI and AI-assisted technologies in the manuscript preparation process

During the preparation of this work, the authors used OpenAI ChatGPT and Anthropic Claude to support language editing, consistency checking, code review and manuscript formatting. After using these tools, the authors reviewed and edited all content and take full responsibility for the published article.

## Notes

### Competing Interest Statement

The authors have declared no competing interest.

### Author Declarations

Statistics Poland Local Data Bank and Copernicus Atmosphere Monitoring Service reanalysis products are publicly available datasets. No IRB or ethics committee approval was required for this analysis of aggregate administrative data.

