## Supplementary Material for "Place-based structural disadvantage and mortality inequality across 379 Polish counties, 2010–2025: a spatial Mundlak analysis"

*[Blinded supplementary file]*

### S1. Supplementary methods

#### S1.1 Conceptual interpretation of structural and within-county layers

The Dahlgren–Whitehead model is used as a heuristic framework for locating measured county determinants within layered influences on population health. It is not treated as a causal DAG. We also distinguish measured exposures from contextual markers rather than labelling every determinant a proxy. PM2.5 is a modelled environmental exposure; education, unemployment, wages, migration, density and peripherality are area indicators that may index broader socioeconomic, compositional or accessibility configurations; physician density indicates provider capacity but not realised access, quality or utilisation. Because health behaviours, psychosocial conditions, social resources and detailed service use are incompletely observed, some associations may partly reflect unmeasured intermediate processes. County-level terms are therefore interpreted as model-conditional markers of linked place domains rather than isolated biological, behavioural or policy mechanisms (Dahlgren and Whitehead, 1991, 2021; Rój and Jankowiak, 2021) Educational composition as a marker of structural disadvantage and its gradient with mortality has been documented across European countries (Mackenbach et al., 2015; Östergren et al., 2017).

#### S1.2 Mortality construction and harmonisation

County deaths and population denominators were obtained from Statistics Poland’s Local Data Bank. Source categories were harmonised to 18 age groups (0–4 through 85+) separately for women and men. Direct age-sex standardisation used one fixed reference distribution pooled across the study counties in 2010–2019. For every county-year, age-sex-specific death rates were multiplied by these fixed weights and summed, then scaled per 100,000. The same weights were applied through 2025.

#### S1.3 Census-derived education series

Comparable education composition was available at the 2011 and 2021 censuses. Higher education shares used the higher-education category divided by the census denominator. Primary/lower-secondary education combined lower-secondary and completed-primary categories in 2011 and used the corresponding combined category in 2021. Annual values were generated by linear interpolation between anchors and linear extrapolation to 2010 and 2022–2025, constrained to 0–100%. Between-county education therefore summarises structural composition, while within-county education represents movement along this constructed trajectory and is not an independently observed annual shock. The two indicators are not complements: upper-secondary and vocational education constitute a substantial intermediate category. Higher-education share therefore captures the upper tail of the county educational distribution, whereas primary/lower-secondary share captures its lower tail. This distinction is conceptually relevant because area higher-education composition can index contextual human capital, whereas educational expansion can make the remaining low-education group increasingly selected for socioeconomic disadvantage (Bor et al., 2024; Östergren et al., 2017).

#### S1.4 PM2.5 construction

PM2.5 was derived from CAMS EAC4 for 2010–2012 and the CAMS European air-quality reanalysis from 2013 onward. EAC4 has a 0.75° global grid and 3-hourly output; the European product has a 0.1° grid and hourly output. European data were sampled every three hours to match EAC4. Product-specific grids were mapped to county polygons separately.

Grid-cell centres inside a county were averaged; for counties without an EAC4 grid centre, the nearest grid point to a representative county point was used. Concentrations were harmonised to μg/m³, averaged spatially by sampling time, then daily, then annually (Copernicus Atmosphere Monitoring Service, 2026a, 2026b).

#### S1.5 Model architecture and basis selection

The final architecture assigned every Mundlak component a national shrinkage smooth and a spatially varying by-smooth, together with a baseline spatial surface and smooth calendar time. Basis dimensions were chosen on Model 1 using a 48-model grid and an additional baseline 175→200 boundary check. Selection targeted the smallest basis producing stable substantive functions under upward increases in k rather than the lowest AIC alone. Concurvity was descriptive, not a fail criterion, because it is structurally expected when spatially patterned global and local terms are estimated jointly. All models were estimated using mgcv::bam with fREML and a gamma inflation parameter of 1.4 (Wood, 2017).

#### S1.6 Strict leave-one-year-out validation

For each omitted year, Mundlak means, lag availability and scaling were reconstructed from training years only. The held-out year therefore did not contribute to preprocessing. The temporal basis was capped only where the training fold had insufficient unique time values; global and spatial basis dimensions remained fixed. This validation targets temporal transportability, which is a harder task than explaining the observed panel and is particularly sensitive to the COVID-19 regime shift.

#### S1.7 Structured-noise perturbation

The perturbation analysis re-exposed the complete fitted GAM to residual-scale noise with empirical temporal and spatial structure using 50 replicates per model. Stability was evaluated continuously using effect-curve correlation, APS sign agreement, signal-to-noise ratio, and—where the fitted local surface was materially heterogeneous—spatial-surface correlation and county sign agreement. The perturbation classification was not used to suppress displayed terms; the full estimates are retained so that robustness and substantive magnitude can be judged separately.

**S1.8 High/low mortality contribution profiles**

Primary profile groups were defined independently of the model as the bottom and top deciles of observed county mean ASMR within 2010–2019, 2020–2022 and 2023–2025. Model 3 was then used to decompose determinant-specific fitted contributions in those outcome-defined groups. A separate sensitivity analysis defined low/high groups by total determinant-only model burden rather than observed mortality; it is shown in Figure S10 to distinguish outcome-defined profiling from decomposition of the model score itself.

**S1.9 Interpretation of within-county components and rationale for Mundlak**

Within-county terms are retained because they answer a distinct temporal question rather than because they are assumed to be causal shocks. A stable within association means that departures from a county's own typical level co-vary reproducibly with next-year mortality conditional on the fitted model. An unstable or sign-reversing association can instead reflect measurement error, selective migration, compositional change, behaviour, psychosocial conditions, disrupted care or another time-varying process that moves with the measured indicator. Such terms are therefore described as conditional temporal coupling or ecological co-movement, not as causal effects. Mundlak separation keeps this short-run layer distinct from persistent structural differences. Interpretation uses four criteria: temporal sign persistence, smooth-shape stability, consistency with the measurement process and substantive plausibility; education-within receives the strongest caveat because its annual path is interpolated/extrapolated from two census anchors (Puchalski and Korzeniowska, 2011).

#### S1.10 Local coefficient-covariance uncertainty for mapped contrasts

For every county-specific marginal or accumulated contrast we propagated model uncertainty through the mgcv linear-predictor matrix. Let  denote the row-vector contrast between the two prediction scenarios for county i and  the fitted coefficient covariance matrix. The model-based standard error is . Approximate pointwise 95% intervals were formed as estimate ±1.96SE and classified as robust negative, uncertain (interval includes zero), or robust positive. For within-county contrasts averaged across years, the linear-predictor contrast was averaged first and its uncertainty propagated from the resulting contrast vector rather than by averaging county-year standard errors. These intervals quantify conditional estimation uncertainty in the fitted GAM. They are not multiplicity-adjusted spatial tests, do not establish causal local effects, and do not replace the structured-noise perturbation analysis. The reporting strategy follows the principle that local SVC point estimates should be shown together with their uncertainty rather than mapped as equally precise everywhere (Mennis, 2006; Irekponor and Oshan, 2025). For determinant-specific contrasts, the focal determinant retained both its national smooth and its corresponding SVC term while non-focal determinant components were set to zero. County locations were retained, so dispersion between county-specific curves represents fitted spatial modification by the focal SVC. IQR and P10–P90 ribbons therefore describe cross-county heterogeneity, not sampling uncertainty; narrow or collapsed ribbons indicate little fitted spatial modification. County-level robust-sign cartograms for Models 1–3 are shown in Supplementary Figures S11–S16; aggregate uncertainty coverage and cross-window surface-stability summaries are shown in Supplementary Figures S17–S18.

**S1.11 Why marginal and accumulated maps can have opposite signs**

Marginal and accumulated maps answer different questions. Suppose a county lies below the 0-SD reference on a non-linear determinant curve and its fitted contribution at the observed level is −15 ASMR per 100,000 relative to 0 SD; the accumulated map is therefore negative. At that same point, however, a +1-SD local increase may move the fitted outcome upward by +20 per 100,000, so the marginal map is positive. There is no contradiction: accumulated contribution asks where the county's current fitted contribution lies relative to the chosen reference, whereas the marginal map asks in which direction the fitted outcome would move from the county's current determinant level. Dominance maps add a second layer of relativity: they identify whichever accumulated contribution is largest within a county, not the determinant with the largest national APS. A determinant with a small national slope can therefore appear as dominant when its observed level is far from the 0-SD reference or when competing contributions are smaller. Contribution sign and map dominance should never be used as universal risk/protective labels. This distinction is particularly important for distance to a city with county rights in Model 3: although distance appears frequently as the largest positive accumulated contribution, its national APS remains small and its determinant-specific spatial modifier is effectively shrunk to zero. That visual dominance therefore reflects a reference-based accumulated contrast and smaller competing contributions, not the emergence of a strong post-COVID distance effect.

**S1.12 Historical-spatial context as interpretive backdrop**

Historical geography is used only to contextualise spatial configurations, not as an unmeasured causal explanation. Poland's former partition territories were exposed to different nineteenth-century institutions and infrastructure, but research finds mixed persistence: some railway, cultural and educational legacies remain, whereas many present-day economic discontinuities do not follow former borders (Grosfeld and Zhuravskaya, 2015; Herbst,

2023). A separate post-war/post-socialist layer is relevant in northern and western areas where state-owned farms were concentrated and whose rapid liquidation after 1991 was followed by persistent unemployment and socioeconomic marginalisation in many localities (Bąk et al., 2021). Polish deprivation research also identifies disadvantaged low-density subregions in eastern and north-western Poland, including overlaps with areas strongly affected by agricultural restructuring (Jamiołkowski et al., 2023). Southern particulate pollution has a different historical-material geography related to solid-fuel heating, industrialisation and topography, especially in Upper Silesia and adjacent southern Poland (Kobza et al., 2018). None of these historical contexts is encoded as an exposure in the fitted GAM, so geographic resemblance is hypothesis-generating rather than evidence of historical causation. County-level labour-market and wage inequality in Poland is documented in OECD (2025).

**S1.13 Same-fit SVC-zeroing sensitivity**

To assess whether determinant-specific SVCs materially alter the reported focal relationships, we performed a post-estimation zeroing sensitivity analysis on each already fitted GAM. For determinant j, the current contrast retains ; the sensitivity contrast sets the focal SVC contribution  to zero while retaining the fitted national smooth . No model was refitted. We compared the current and SVC-zeroed APS, national sign, county marginal-map sign agreement and accumulated-map sign agreement. This check distinguishes spatial modification of magnitude or local pattern from a change in the overall national direction (Supplementary Table S10).

**S1.14 Interpreting cross-window sign changes and post-COVID reweighting**

Models 1–3 are overlapping analytical windows rather than separate era-specific fits, so APS differences across them are sensitivity comparisons and not clean pre-/post-COVID causal contrasts. The within-county layer is especially regime-sensitive: PM2.5 changes from approximately zero in Model 1 to +4.6 in Model 2 and −13.2 in Model 3; higher education changes from approximately zero to −8.7 and then +2.0; long-term unemployment becomes more negative in Models 2–3. These reversals do not imply that established risk factors became protective. They indicate altered temporal coupling after 2020. Plausible contributors include pandemic-era co-movement of employment, mobility, energy use and health-care utilisation; selective mortality or mortality displacement among frailer populations; secular improvement and compressed within-country variability in exposures such as PM2.5; and a lower signal-to-noise ratio when remaining temporal variation is small relative to measurement error, shared trends and residual confounding. These mechanisms are not separately identified by the present ecological models. The outcome-defined HIGH–LOW profiles provide a complementary period-specific description: education remains a strong separator after COVID, whereas the PM2.5 contribution contrast attenuates sharply. This is interpreted as reweighting of a multidimensional place profile rather than disappearance of the underlying structural gradient. Long-term unemployment illustrates the same distinction: its persistent between-county gradient remains positive, while its within-county association becomes more negative in the pandemic-inclusive windows. This does not imply that unemployment became protective; short-run unemployment deviations may instead have co-moved with pandemic labour-market interventions, mobility, sectoral composition, migration and other unmeasured changes. Likewise, a secular decline in PM2.5 does not by itself generate a negative within slope; rather, lower pollution during periods of unusually high pandemic mortality, together with reduced within-country exposure variability, can contribute to an inverse temporal association after conditioning on the other model components.

**Supplementary Table S1.** Measured determinants, Dahlgren–Whitehead layer, model role and interpretive caveat.

| Determinant | Conceptual layer | Mundlak role | Interpretive caveat |
|---|---|---|---|
| Higher education | Living/working conditions; broader socioeconomic context | Between + lag-1 within | Census-derived structural socioeconomic and area-level human-capital marker; within trajectory is interpolated/extrapolated. May index professional employment, institutional resources, health literacy, social networks, selective migration and health-related norms; not an isolated schooling effect. |
| Primary/lower-secondary education | Living/working conditions; broader socioeconomic context | Between + lag-1 within | Census-derived marker of the lower tail of the educational distribution and accumulated socioeconomic disadvantage. Same within-trajectory limitation; may co-vary with labour-market exclusion, limited socioeconomic mobility, health literacy, health-related behaviours and selective migration. It is not the algebraic complement of higher education because upper-secondary and vocational education form a substantial intermediate category. |
| Long-term unemployment | Living/working conditions | Between + lag-1 within | Marker of labour-market exclusion rather than a self-contained exposure. May co-vary with income insecurity, inactivity, selective migration, local restructuring and psychosocial disadvantage. |
| Gross wages | Living/working conditions / socioeconomic resources | Between + lag-1 within | Area-level marker of local earnings, productivity and occupational composition; does not equal household disposable income or individual material resources. |
| Physicians per 10,000 | Health-care services | Between + lag-1 within | Indicator of local provider capacity/service concentration; does not measure realised access, quality, continuity or utilisation and is partly patterned by urban–rural workforce distribution. |
| PM2.5 | General environmental conditions | Between + lag-1 within | Modelled environmental exposure, not primarily a proxy. Spatial pattern can reflect domestic solid-fuel heating, industry, transport and topography; 2013 product-resolution change and pandemic co-movement limit short-run interpretation. |
| Net migration per 1,000 | Broader socioeconomic context / social composition | Between + lag-1 within | Marker of demographic sorting and local attractiveness; may index age/skill composition, labour and housing opportunities rather than an exogenous migration shock. |
| Population density | Living conditions / urban context | Between + lag-1 within | Composite urbanicity marker that may bundle service concentration, labour-market access, housing, transport, social composition and environmental exposures. |
| Distance to city with county rights | Living conditions / access and peripherality | Between only | Accessibility/peripherality marker; does not directly measure travel time, transport quality, actual service use or economic connectivity. |

**Supplementary Table S2.** Data provenance and construction for final model variables.

| Measure | Source | Identifier/product | Construction |
|---|---|---|---|
| ASMR | Statistics Poland BDL death/population tables | Deaths: P1343 dictionary; population: P2137 dictionary | 18 age groups × 2 sexes; fixed 2010–2019 pooled standard; per 100,000 |
| Higher education (%) | Statistics Poland population censuses | 2011: 396203 / 396216; 2021: 1701451 / 1701450 | Annual linear interpolation/extrapolation, bounded 0–100% |
| Primary/lower-secondary education (%) | Statistics Poland population censuses | 2011: (396139 + 396126) / 396216; 2021: 1701456 / 1701450 | Annual linear interpolation/extrapolation, bounded 0–100% |
| Long-term unemployment (%) | Statistics Poland BDL | 79227 | County-year value |
| Gross wages (PLN) | Statistics Poland BDL | 64428 | County-year value |
| Physicians per 10,000 | Statistics Poland BDL | 454186 | County-year value |
| Net migration per 1,000 | Statistics Poland BDL | 453193 | County-year value |
| Population density (persons/km²) | Population + county GIS area | Derived | Annual population divided by county polygon area in km² |
| Distance to city with county rights (km) | County GIS geometry | Derived | Centroid-to-nearest city-with-county-rights distance; city counties set to 0 |
| Annual mean PM2.5 (μg/m³) | CAMS EAC4 / CAMS European air-quality reanalysis | EAC4 DOI 10.24381/d58bbf47; Europe DOI 10.24381/7cc0465a | 2010–2012 EAC4; 2013–2025 Europe; 3-hourly harmonisation; county spatial/daily/annual averaging |

**Supplementary Table S3.** Basis-dimension sensitivity and retained specification. "Basis" denotes the maximum basis dimension (k) allowed for the corresponding smooth; the table reports stability when additional flexibility was allowed.

| **Dimension** | **Increase** | **Median ΔAIC** | **Median ΔBIC** | **APS corr.** | **P10 surface corr.** | **P10 curve corr.** | **Stability pass** |
|---|---|---|---|---|---|---|---|
| National determinant smooth basis | 3->5 | -9.9 | 11.0 | 0.9941 | 0.9969 | 0.9963 | 79% |
| Spatial modifier basis | 25->35 | -11.8 | 41.9 | 0.9993 | 0.9958 | 0.9999 | 75% |
| Spatial modifier basis | 35->45 | -5.2 | 1.5 | 0.9996 | 0.9973 | 1.0000 | 75% |
| Baseline spatial smooth basis | 100->125 | -93.6 | 3.3 | 0.9888 | 0.7566 | 0.9993 | 0% |
| Baseline spatial smooth basis | 125->150 | -65.5 | 30.4 | 0.9899 | 0.9818 | 0.9962 | 50% |
| Baseline spatial smooth basis | 150->175 | -22.2 | 13.8 | 0.9864 | 0.9934 | 0.9974 | 33% |
| Calendar-time smooth basis | 7->9 | -128.4 | -148.4 | 0.9963 | 0.9913 | 0.9997 | 79% |
| Baseline spatial smooth basis | 175→200 | −5.4 | +34.3 | 0.9995 | 0.9988 | 0.9991 | 100% |

Retained specification: basis dimension 5 for determinant-specific national smooths, 25 for determinant-specific spatially varying coefficient surfaces, 175 for the baseline spatial smooth, and 9 for calendar time. The 175→200 boundary refit had APS sign agreement 1.00, mean county sign agreement 0.990, ΔRMSE −0.13, ΔAIC −5.44 and ΔBIC +34.30; the smaller baseline basis was therefore retained.

**Supplementary Table S4.** Extended fit, residual and strict temporal-validation diagnostics for the three fitted analytical windows.

| Period | n | $R^2$ | Dev. expl. | RMSE | AIC | BIC | Median Moran I | Positive Moran $p<.05$ | Median ACF1 | $\|ACF1\|>.2$ | LOO median $R^2$ | LOO mean RMSE |
|---|---|---|---|---|---|---|---|---|---|---|---|---|
| 2010–2019 | 3,411 | 0.731 | 0.744 | 49.8 | 36707.4 | 37818.7 | −0.025 | 0/9 | −0.025 | 58.3% | 0.479 | 77.0 |
| 2010–2022 | 4,548 | 0.803 | 0.811 | 53.4 | 49511.8 | 50873.1 | +0.008 | 1/12 | −0.046 | 51.5% | −3.544 | 568.4 |
| 2010–2025 | 5,685 | 0.787 | 0.794 | 57.8 | 62700.5 | 64136.4 | +0.025 | 4/15 | −0.030 | 50.9% | 0.326 | 455.2 |

**Supplementary Table S5.** Linear Mundlak ridge benchmark across analytical windows: debiased coefficient [95% dependent-multiplier bootstrap interval].

| **Determinant** | **Layer** | **2010–2019** | **2010–2022** | **2010–2025** |
|---|---|---|---|---|
| Higher education | Between | −9.13 [−10.04, −8.20] | −10.11 [−11.03, −9.30] | −9.83 [−10.59, −9.10] |
| Primary / lower-secondary education | Between | +7.22 [+6.33, +8.12] | +7.15 [+6.30, +7.96] | +6.29 [+5.64, +6.99] |
| Long-term unemployment | Between | +3.43 [+2.44, +4.42] | +3.53 [+2.56, +4.54] | +3.04 [+2.17, +3.94] |
| Gross wages | Between | −2.73 [−3.66, −1.69] | −2.68 [−3.60, −1.80] | −2.32 [−3.24, −1.45] |
| Physicians | Between | −6.99 [−8.09, −5.89] | −7.51 [−8.54, −6.45] | −7.88 [−8.84, −6.99] |
| PM2.5 | Between | +3.75 [+2.37, +5.07] | +3.33 [+2.09, +4.59] | +2.83 [+1.78, +3.99] |
| Net migration | Between | −3.24 [−4.27, −2.22] | −4.17 [−5.20, −3.14] | −2.99 [−3.95, −2.08] |
| Population density | Between | −2.45 [−3.91, −0.91] | −2.36 [−3.64, −1.02] | −2.49 [−3.75, −1.34] |
| Distance to city with county rights | Between | +4.21 [+3.10, +5.30] | +4.22 [+3.26, +5.23] | +4.12 [+3.28, +5.01] |
| Higher education | Within | +0.41 [+0.13, +0.70] | +0.15 [−0.23, +0.51] | +0.57 [+0.18, +0.95] |
| Primary / lower-secondary education | Within | +0.43 [+0.13, +0.74] | +0.44 [≈0, +0.83] | +0.70 [+0.32, +1.09] |
| Long-term unemployment | Within | −1.47 [−2.36, −0.65] | −2.57 [−3.40, −1.77] | −2.11 [−2.81, −1.39] |
| Gross wages | Within | +0.33 [≈0, +0.68] | −1.58 [−2.02, −1.11] | −1.34 [−2.00, −0.75] |
| Physicians | Within | +0.31 [−0.76, +1.42] | −2.43 [−3.55, −1.32] | −0.98 [−2.05, ≈0] |
| PM2.5 | Within | +2.34 [+1.30, +3.41] | −1.00 [−1.93, −0.09] | −0.65 [−1.44, +0.15] |
| Net migration | Within | +1.01 [+0.05, +1.93] | +0.50 [−0.46, +1.51] | +0.78 [−0.10, +1.71] |
| Population density | Within | +2.05 [+0.40, +3.66] | +0.77 [−0.83, +2.47] | +1.15 [−0.22, +2.71] |

Intervals are from the recentered dependent-multiplier bootstrap. The ridge model is a homogeneous linear benchmark and is not used to select the GAM.

**Supplementary Table S6.** Structured-noise perturbation summaries across analytical windows (50 refits per model).

| Determinant | Layer | 2010–2019 | 2010–2022 | 2010–2025 |
|---|---|---|---|---|
| Higher education | Between | SNR 38.7; r 1.000; APS sign 100%; surf 1.00 | SNR 37.2; r 1.000; APS sign 100%; surf 0.98 | SNR 33.0; r 0.999; APS sign 100%; surf 0.99 |
| Primary / lower-secondary education | Between | SNR 38.1; r 0.999; APS sign 100%; surf 0.96 | SNR 34.5; r 0.998; APS sign 100%; surf 0.86 | SNR 30.4; r 0.996; APS sign 100%; surf 0.87 |
| Long-term unemployment | Between | SNR 19.4; r 0.973; APS sign 100%; surf 0.93 | SNR 22.1; r 0.956; APS sign 100%; surf 0.98 | SNR 20.6; r 0.995; APS sign 100%; surf 0.92 |
| Gross wages | Between | SNR 11.0; r 0.995; APS sign 100%; surf 0.98 | SNR 2.5; r 0.743; APS sign 86%; surf 0.95 | SNR 3.1; r 0.927; APS sign 100%; surf 0.89 |
| Physicians | Between | SNR 1.7; r 0.760; APS sign 88%; surf 0.88 | SNR 11.7; r 0.681; APS sign 98%; surf 0.98 | SNR 18.1; r 0.999; APS sign 100%; surf 0.98 |
| PM2.5 | Between | SNR 27.8; r 1.000; APS sign 100%; surf 0.95 | SNR 26.2; r 1.000; APS sign 100%; surf 0.95 | SNR 30.0; r 0.995; APS sign 100%; surf 0.93 |
| Net migration | Between | SNR 0.0; r 1.000; APS sign 100%; surf NA | SNR 0.3; r -0.240; APS sign 38%; surf NA | SNR 0.0; r 0.840; APS sign 92%; surf NA |
| Population density | Between | SNR 38.2; r 1.000; APS sign 100%; surf NA | SNR 33.1; r 1.000; APS sign 100%; surf NA | SNR 2.4; r 1.000; APS sign 100%; surf 0.92 |
| Distance to city with county rights | Between | SNR 0.0; r 1.000; APS sign 100%; surf NA | SNR 0.0; r 0.332; APS sign 66%; surf NA | SNR 8.7; r 0.026; APS sign 16%; surf NA |
| Higher education | Within | SNR 0.0; r 0.242; APS sign 62%; surf NA | SNR 20.5; r 0.985; APS sign 100%; surf NA | SNR 1.7; r 0.755; APS sign 86%; surf 0.96 |
| Primary / lower-secondary education | Within | SNR 5.8; r 0.905; APS sign 100%; surf 0.99 | SNR 5.8; r 0.819; APS sign 98%; surf 0.92 | SNR 19.6; r 1.000; APS sign 100%; surf 0.97 |
| Long-term unemployment | Within | SNR 5.0; r 1.000; APS sign 100%; surf NA | SNR 27.9; r 1.000; APS sign 100%; surf 0.95 | SNR 12.3; r 0.987; APS sign 100%; surf 0.99 |
| Gross wages | Within | SNR 0.0; r 0.999; APS sign 100%; surf NA | SNR 1.7; r 0.642; APS sign 86%; surf 0.94 | SNR 4.1; r 0.906; APS sign 100%; surf 0.98 |
| Physicians | Within | SNR 0.3; r 0.560; APS sign 78%; surf NA | SNR 5.1; r 1.000; APS sign 100%; surf 0.93 | SNR 0.0; r 0.000; APS sign 50%; surf NA |
| PM2.5 | Within | SNR 0.0; r 0.755; APS sign 88%; surf NA | SNR 7.8; r 0.913; APS sign 100%; surf 0.96 | SNR 56.6; r 1.000; APS sign 100%; surf 0.97 |
| Net migration | Within | SNR 0.0; r 0.920; APS sign 96%; surf NA | SNR 5.4; r 0.960; APS sign 98%; surf 0.96 | SNR 8.1; r 0.955; APS sign 100%; surf 0.97 |
| Population density | Within | SNR 29.1; r 1.000; APS sign 100%; surf NA | SNR 8.5; r 1.000; APS sign 100%; surf NA | SNR 14.7; r 1.000; APS sign 100%; surf NA |

SNR = baseline curve range divided by perturbation-induced curve variability; r = mean curve-shape correlation; APS sign = percentage of perturbation refits preserving the baseline APS sign. "Spatial r (P10)" refers to the 10th percentile of county-surface correlations and is interpreted only when the spatial surface has non-negligible variation. These are sensitivity diagnostics, not confidence intervals or pass/fail significance tests.

**Supplementary Table S7.** Raw determinant profiles in counties with LOW versus HIGH observed mortality in the three descriptive periods: median [IQR]. LOW/HIGH are bottom/top deciles of observed county mean ASMR within each period.

| **Determinant** | **Unit** | **2010–2019 LOW** | **2010–2019 HIGH** | **COVID 2020–2022 LOW** | **COVID 2020–2022 HIGH** | **Post-COVID 2023–2025 LOW** | **Post-COVID 2023–2025 HIGH** |
|---|---|---|---|---|---|---|---|
| Higher education | % | 24.3 [16.7–30.7] | 14.1 [12.3–15.1] | 28.8 [23.4–35.8] | 16.7 [15.7–18.0] | 29.8 [20.8–36.9] | 19.3 [17.9–20.8] |
| Primary / lower-secondary education | % | 14.5 [12.7–20.2] | 23.9 [21.0–26.9] | 11.2 [9.7–13.8] | 18.3 [15.8–20.6] | 10.2 [8.4–13.4] | 14.5 [13.4–16.5] |
| Long-term unemployment | % of economically active | 3.6 [2.0–4.8] | 5.0 [3.9–6.0] | 1.9 [0.9–2.8] | 3.8 [2.3–5.7] | 1.7 [1.0–2.6] | 2.4 [1.7–3.7] |
| Gross wages | PLN | 3 712 [3 374–4 145] | 3 366 [3 216–3 599] | 5 524 [5 082–6 105] | 5 062 [4 907–5 288] | 7 354 [7 041–8 441] | 7 256 [7 029–7 564] |
| Physicians | per 10,000 | 35.7 [17.5–48.0] | 13.4 [8.8–17.8] | 38.4 [16.9–69.2] | 15.6 [10.6–19.0] | 40.2 [20.2–79.0] | 17.1 [11.3–20.5] |
| PM2.5 | µg/m³ | 15.7 [13.5–16.5] | 17.1 [15.5–17.8] | 15.7 [13.4–16.7] | 14.6 [13.6–16.0] | 12.6 [11.6–14.0] | 12.6 [11.6–13.8] |
| Net migration | per 1,000 | -1.2 [-3.1–0.8] | -2.0 [-3.1–-0.4] | -1.2 [-2.7–2.9] | -2.6 [-3.5–-2.3] | -2.3 [-3.5–-0.4] | -1.4 [-2.4–0.9] |
| Population density | persons/km² | 1 091 [148–1 904] | 80 [64–193] | 847 [154–1 827] | 67 [56–103] | 923 [131–1 806] | 63 [50–106] |
| Distance to city with county rights | km | 0.0 [0.0–19.0] | 27.7 [1.7–39.0] | 0.0 [0.0–16.7] | 32.9 [19.5–46.6] | 0.0 [0.0–24.8] | 31.6 [19.5–47.8] |

**Supplementary Table S8.** HIGH-minus-LOW mean Model-3 determinant contribution in observed-mortality groups across the three descriptive periods (ASMR per 100,000).

| Determinant | Layer | 2010–2019 | COVID 2020–2022 | Post-COVID 2023–2025 |
|---|---|---|---|---|
| Higher education | Between | +68.9 | +79.3 | +58.5 |
| Primary / lower-secondary education | Between | +58.0 | +52.0 | +48.6 |
| Long-term unemployment | Between | +5.3 | +9.5 | +4.1 |
| Gross wages | Between | +4.0 | +4.9 | +9.2 |
| Physicians | Between | −23.0 | −19.9 | −23.2 |
| PM2.5 | Between | +22.1 | +8.2 | +0.3 |
| Net migration | Between | ≈0 | ≈0 | ≈0 |
| Population density | Between | +1.5 | +1.1 | +0.8 |
| Distance to city with county rights | Between | −5.6 | −5.1 | −6.6 |
| Higher education | Within | +3.0 | −2.6 | −8.8 |
| Primary / lower-secondary education | Within | +4.1 | −1.0 | +3.5 |
| Long-term unemployment | Within | −0.4 | +0.1 | −3.7 |
| Gross wages | Within | −4.8 | +1.4 | +15.5 |
| Physicians | Within | ≈0 | ≈0 | ≈0 |
| PM2.5 | Within | +0.5 | +5.5 | −1.7 |
| Net migration | Within | +0.3 | +1.8 | +0.8 |
| Population density | Within | +0.7 | −1.0 | +0.3 |

Positive values mean that the determinant term made a more positive (or less negative) fitted contribution in the HIGH observed-ASMR group than in the LOW group. These are reference-based contribution contrasts, not marginal slopes or causal attributable effects. The sharp attenuation of the PM2.5 contrast from pre-COVID to 2023–2025 therefore indicates reduced separation of the outcome-defined groups by this fitted contribution, not disappearance of the positive between-county PM2.5 APS.

**Supplementary Table S9.** Local robust-sign coverage for selected Model 1 contrasts (% of 379 counties).

| Determinant | Layer | Map | Robust positive | Robust negative | Uncertain |
|---|---|---|---|---|---|
| Higher education | Between | Marginal | 0.5 | 65.7 | 33.8 |
| Primary/lower-secondary education | Between | Marginal | 55.4 | 5.5 | 39.1 |
| Long-term unemployment | Between | Marginal | 31.9 | 0.0 | 68.1 |
| PM2.5 | Between | Marginal | 72.3 | 0.0 | 27.7 |
| Population density | Between | Marginal | 100.0 | 0.0 | 0.0 |
| Physicians | Between | Marginal | 0.0 | 0.0 | 100.0 |
| Net migration | Between | Marginal | 0.0 | 0.0 | 100.0 |
| Distance to city with county rights | Between | Marginal | 0.0 | 0.0 | 100.0 |
| Primary/lower-secondary education | Within | Marginal | 28.2 | 0.0 | 71.8 |
| Population density | Within | Marginal | 100.0 | 0.0 | 0.0 |
| Higher education | Within | Marginal | 0.0 | 0.0 | 100.0 |
| PM2.5 | Within | Marginal | 0.0 | 0.0 | 100.0 |

**Supplementary Table S10.** Same-fit determinant-specific spatial-modifier zeroing sensitivity across Models 1–3. “Spatial modifier EDF” is the effective degrees of freedom of the focal spatially varying coefficient term. “Full focal effect” retains the national smooth plus that spatial modifier; “modifier removed” sets only the focal spatial modifier to zero post-estimation while retaining the fitted national smooth and all other model components. Map agreement is the percentage of counties retaining the same sign after this post-estimation removal. No alternative model was refitted.

| Model | Determinant | Layer | Spatial modifier EDF | APS, full focal effect | APS, modifier removed | Difference in APS | APS sign changed? | Marginal-map sign agreement | Accumulated-map sign agreement |
|---|---|---|---|---|---|---|---|---|---|
| 2010–2019 | Higher education | Between | 2.43 | −26.14 | −10.72 | −15.42 | No | 83.9% | 83.9% |
| 2010–2019 | Primary / lower-secondary education | Between | 15.04 | +29.36 | +8.52 | +20.84 | No | 76.0% | 73.4% |
| 2010–2019 | Long-term unemployment | Between | 0.80 | +7.53 | +7.55 | ≈0 | No | 99.7% | 100.0% |
| 2010–2019 | Gross wages | Between | 2.52 | −2.60 | ≈0 | −2.60 | No | 63.3% | 63.3% |
| 2010–2019 | Physicians | Between | 0.37 | +0.16 | ≈0 | +0.16 | No | 59.4% | 59.4% |
| 2010–2019 | PM2.5 | Between | 1.72 | +30.18 | +22.97 | +7.21 | No | 100.0% | 100.0% |
| 2010–2019 | Net migration | Between | 0.00 | ≈0 | ≈0 | ≈0 | No | 100.0% | 100.0% |
| 2010–2019 | Population density | Between | 0.00 | +15.06 | +15.06 | ≈0 | No | 100.0% | 100.0% |
| 2010–2019 | Distance to city with county rights | Between | 0.00 | ≈0 | ≈0 | ≈0 | No | 100.0% | 100.0% |
| 2010–2019 | Higher education | Within | 0.00 | ≈0 | ≈0 | ≈0 | No | 52.5% | 52.5% |
| 2010–2019 | Primary / lower-secondary education | Within | 1.88 | +7.12 | +6.13 | +0.99 | No | 93.1% | 94.2% |
| 2010–2019 | Long-term unemployment | Within | 0.00 | −1.05 | −1.05 | ≈0 | No | 100.0% | 100.0% |
| 2010–2019 | Gross wages | Within | 0.00 | ≈0 | ≈0 | ≈0 | No | 100.0% | 100.0% |
| 2010–2019 | Physicians | Within | 0.07 | ≈0 | ≈0 | ≈0 | No | 54.6% | 54.6% |
| 2010–2019 | PM2.5 | Within | 0.00 | ≈0 | ≈0 | ≈0 | No | 55.4% | 55.4% |
| 2010–2019 | Net migration | Within | 0.00 | ≈0 | ≈0 | ≈0 | No | 94.7% | 94.7% |
| 2010–2019 | Population density | Within | 0.00 | +3.72 | +3.72 | ≈0 | No | 100.0% | 100.0% |
| 2010–2022 | Higher education | Between | 2.27 | −24.99 | −14.06 | −10.93 | No | 88.1% | 88.4% |
| 2010–2022 | Primary / lower-secondary education | Between | 17.20 | +24.19 | +11.78 | +12.41 | No | 83.1% | 73.4% |
| 2010–2022 | Long-term unemployment | Between | 1.25 | +8.88 | +8.51 | +0.37 | No | 96.3% | 98.7% |
| 2010–2022 | Gross wages | Between | 2.40 | −1.38 | ≈0 | −1.38 | No | 57.5% | 57.5% |
| 2010–2022 | Physicians | Between | 1.70 | +2.83 | +2.99 | −0.16 | No | 55.4% | 53.3% |
| 2010–2022 | PM2.5 | Between | 1.64 | +31.62 | +27.03 | +4.59 | No | 100.0% | 100.0% |
| 2010–2022 | Net migration | Between | 0.00 | ≈0 | ≈0 | ≈0 | No | 100.0% | 100.0% |
| 2010–2022 | Population density | Between | 0.00 | +14.11 | +14.11 | ≈0 | No | 100.0% | 100.0% |
| 2010–2022 | Distance to city with county rights | Between | 0.00 | ≈0 | ≈0 | ≈0 | No | 87.1% | 88.7% |
| 2010–2022 | Higher education | Within | 0.00 | −8.71 | −8.71 | ≈0 | No | 100.0% | 100.0% |
| 2010–2022 | Primary / lower-secondary education | Within | 1.75 | +4.38 | +3.67 | +0.71 | No | 68.3% | 71.5% |
| 2010–2022 | Long-term unemployment | Within | 0.99 | −5.18 | −4.84 | −0.34 | No | 100.0% | 100.0% |

| | | | | | | | | | |
|---|---|---|---|---|---|---|---|---|---|
| 2010–2022 | Gross wages | Within | 1.46 | −0.47 | ≈0 | −0.47 | No | 58.0% | 58.0% |
| 2010–2022 | Physicians | Within | 1.14 | +0.34 | ≈0 | +0.34 | No | 69.7% | 69.7% |
| 2010–2022 | PM2.5 | Within | 1.70 | +4.61 | +4.55 | +0.06 | No | 79.4% | 54.1% |
| 2010–2022 | Net migration | Within | 1.32 | +0.51 | ≈0 | +0.51 | No | 69.9% | 69.9% |
| 2010–2022 | Population density | Within | 0.00 | +0.62 | +0.62 | ≈0 | No | 100.0% | 100.0% |
| 2010–2025 | Higher education | Between | 1.75 | −33.53 | −33.04 | −0.50 | No | 100.0% | 100.0% |
| 2010–2025 | Primary / lower-secondary education | Between | 14.68 | +19.19 | +9.70 | +9.49 | No | 82.6% | 69.1% |
| 2010–2025 | Long-term unemployment | Between | 0.20 | +9.63 | +9.54 | +0.09 | No | 100.0% | 100.0% |
| 2010–2025 | Gross wages | Between | 1.44 | −3.46 | −3.37 | −0.09 | No | 55.7% | 56.2% |
| 2010–2025 | Physicians | Between | 2.20 | +9.90 | +6.24 | +3.66 | No | 84.4% | 84.4% |
| 2010–2025 | PM2.5 | Between | 1.78 | +36.73 | +38.00 | −1.27 | No | 88.1% | 86.3% |
| 2010–2025 | Net migration | Between | 0.00 | ≈0 | ≈0 | ≈0 | No | 74.9% | 74.7% |
| 2010–2025 | Population density | Between | 1.54 | +2.46 | ≈0 | +2.46 | No | 71.8% | 71.8% |
| 2010–2025 | Distance to city with county rights | Between | 0.00 | +0.69 | +0.69 | ≈0 | No | 100.0% | 100.0% |
| 2010–2025 | Higher education | Within | 1.59 | +2.03 | +0.10 | +1.93 | No | 69.9% | 69.9% |
| 2010–2025 | Primary / lower-secondary education | Within | 2.32 | +7.04 | +2.48 | +4.55 | No | 82.6% | 82.6% |
| 2010–2025 | Long-term unemployment | Within | 1.84 | −5.48 | −4.50 | −0.98 | No | 94.7% | 94.2% |
| 2010–2025 | Gross wages | Within | 2.23 | −1.10 | ≈0 | −1.10 | No | 63.6% | 63.6% |
| 2010–2025 | Physicians | Within | 0.00 | ≈0 | ≈0 | ≈0 | No | 50.9% | 50.9% |
| 2010–2025 | PM2.5 | Within | 1.65 | −13.18 | −12.59 | −0.59 | No | 100.0% | 100.0% |
| 2010–2025 | Net migration | Within | 1.31 | +1.32 | +1.17 | +0.16 | No | 89.4% | 86.8% |
| 2010–2025 | Population density | Within | 0.00 | +0.85 | +0.85 | ≈0 | No | 100.0% | 100.0% |

Note. Robust sign means the approximate pointwise 95% coefficient-covariance interval excludes zero. Classification is descriptive and not multiplicity-adjusted. Full percentages for Models 1–3, marginal and accumulated contrasts are provided in the accompanying machine-readable supplementary table.

### S2. Supplementary figures

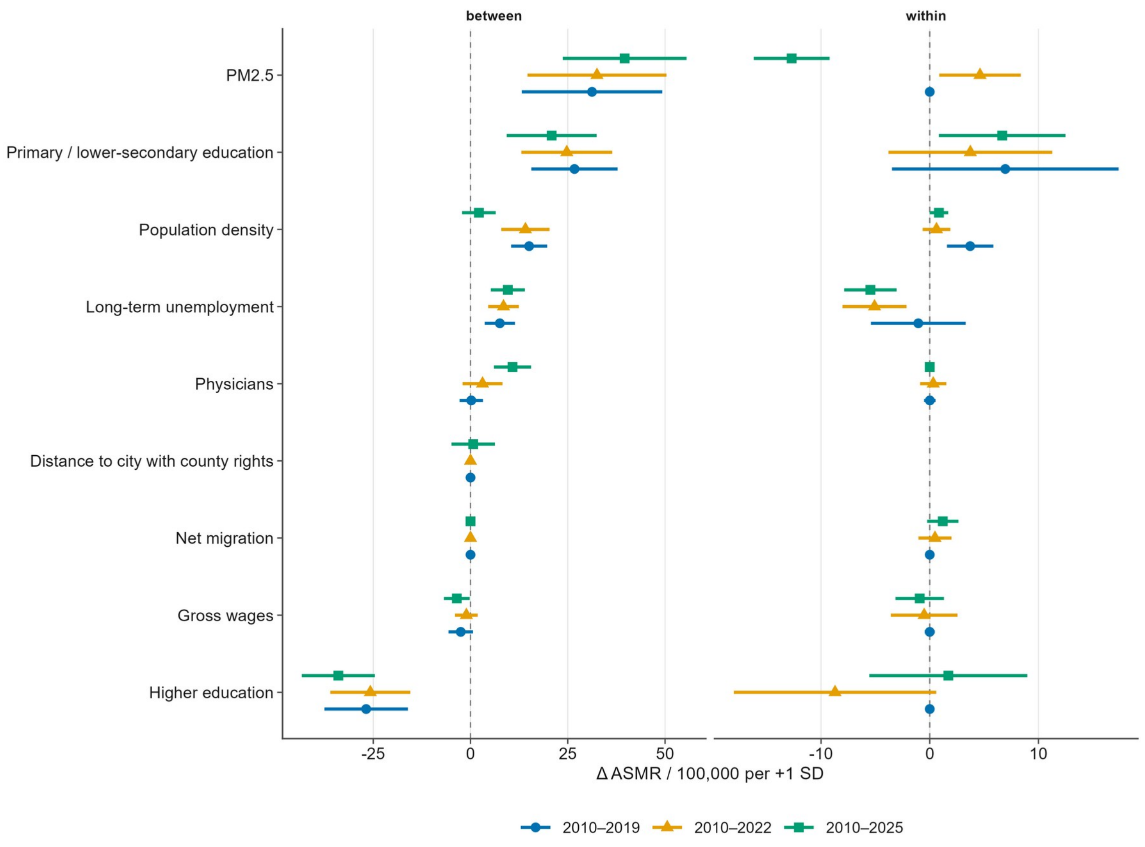


**Supplementary Figure S1.** Cross-period comparison of GAM average partial slopes.

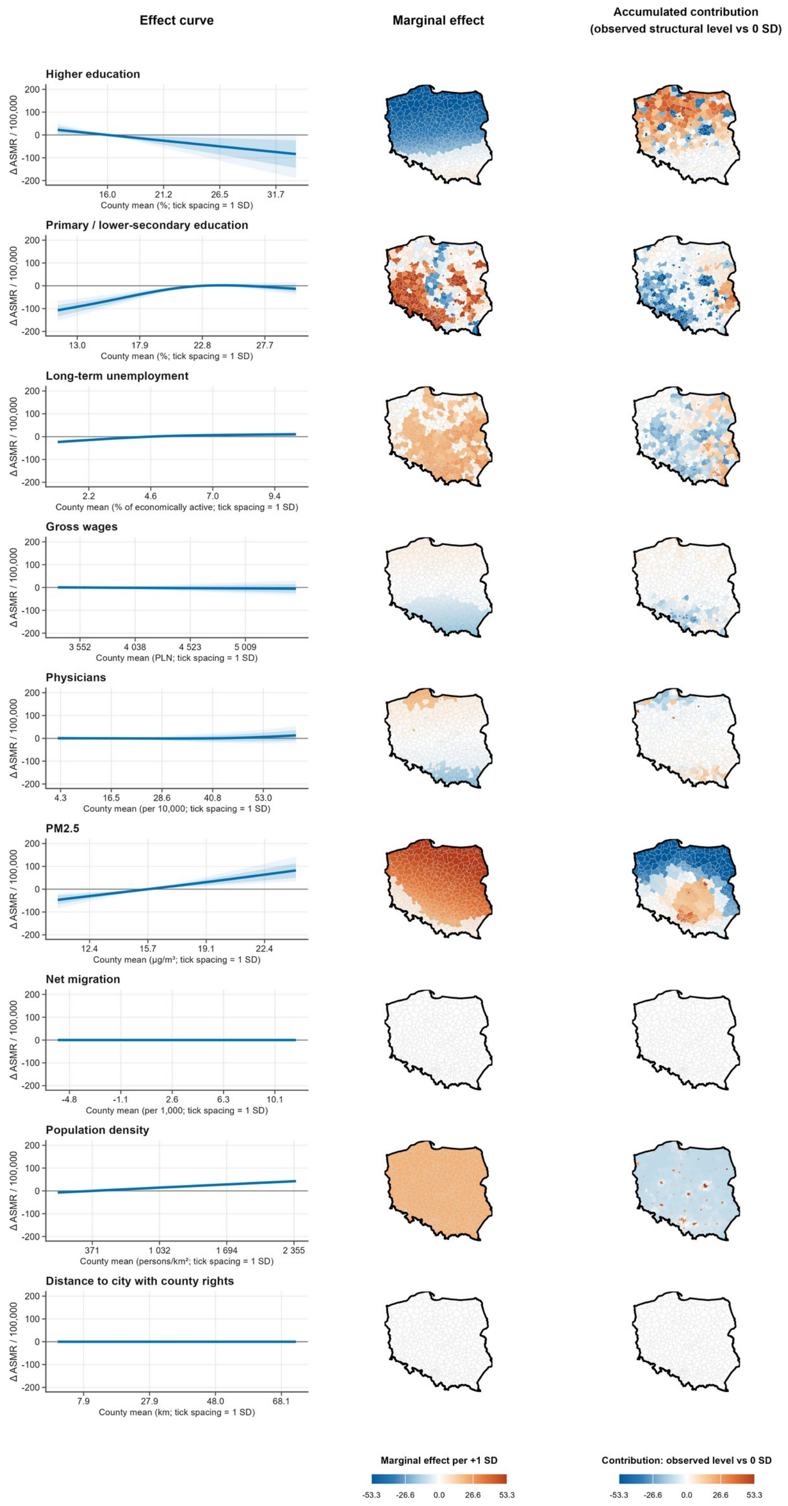


**Supplementary Figure S2.** Full Model 2 between-county effect grid, 2010–2022. Map lines delineate study areas and do not necessarily depict accepted national boundaries.

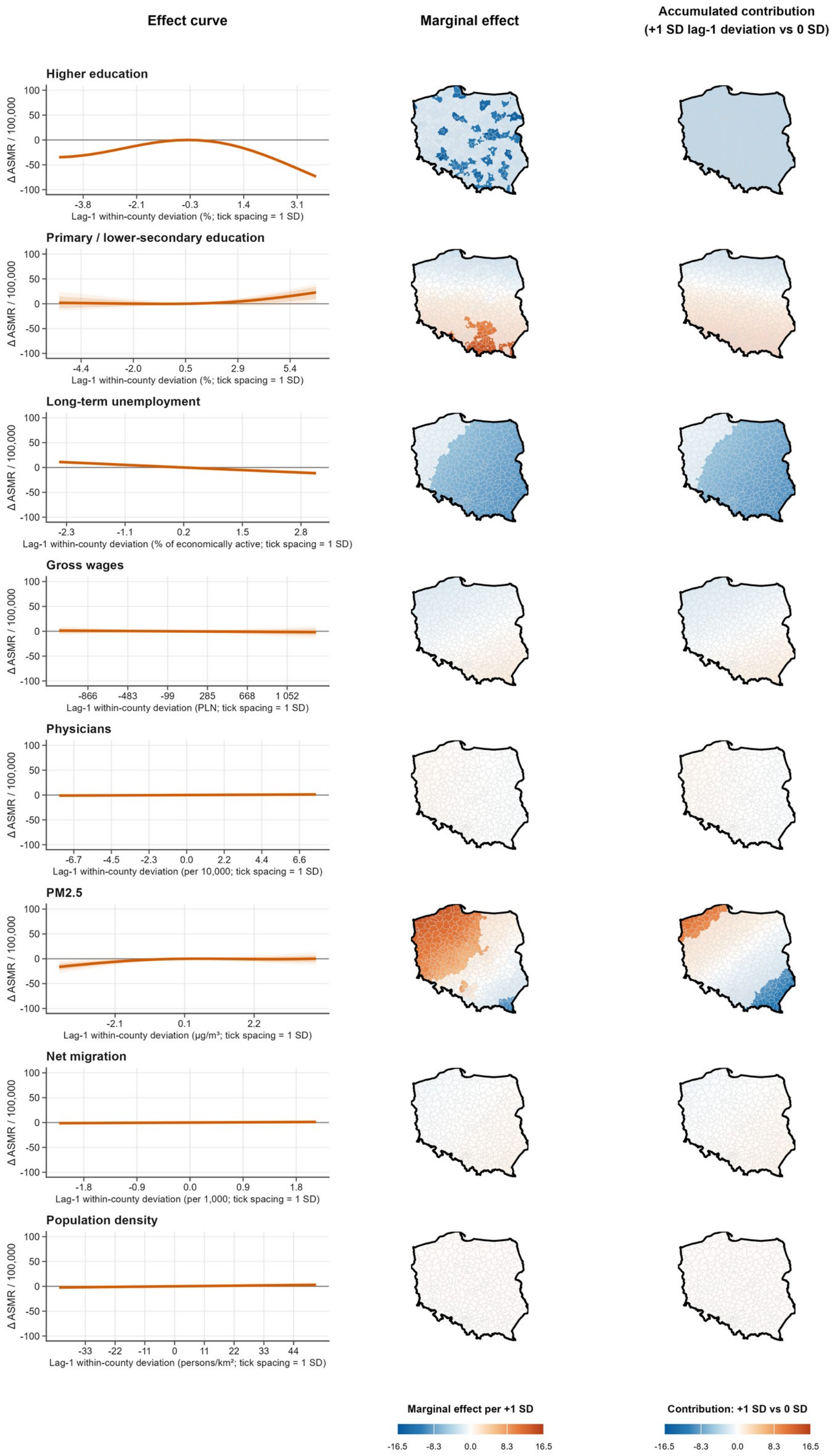


**Supplementary Figure S3.** Full Model 2 within-county lag-1 effect grid, 2010–2022. Map lines delineate study areas and do not necessarily depict accepted national boundaries.

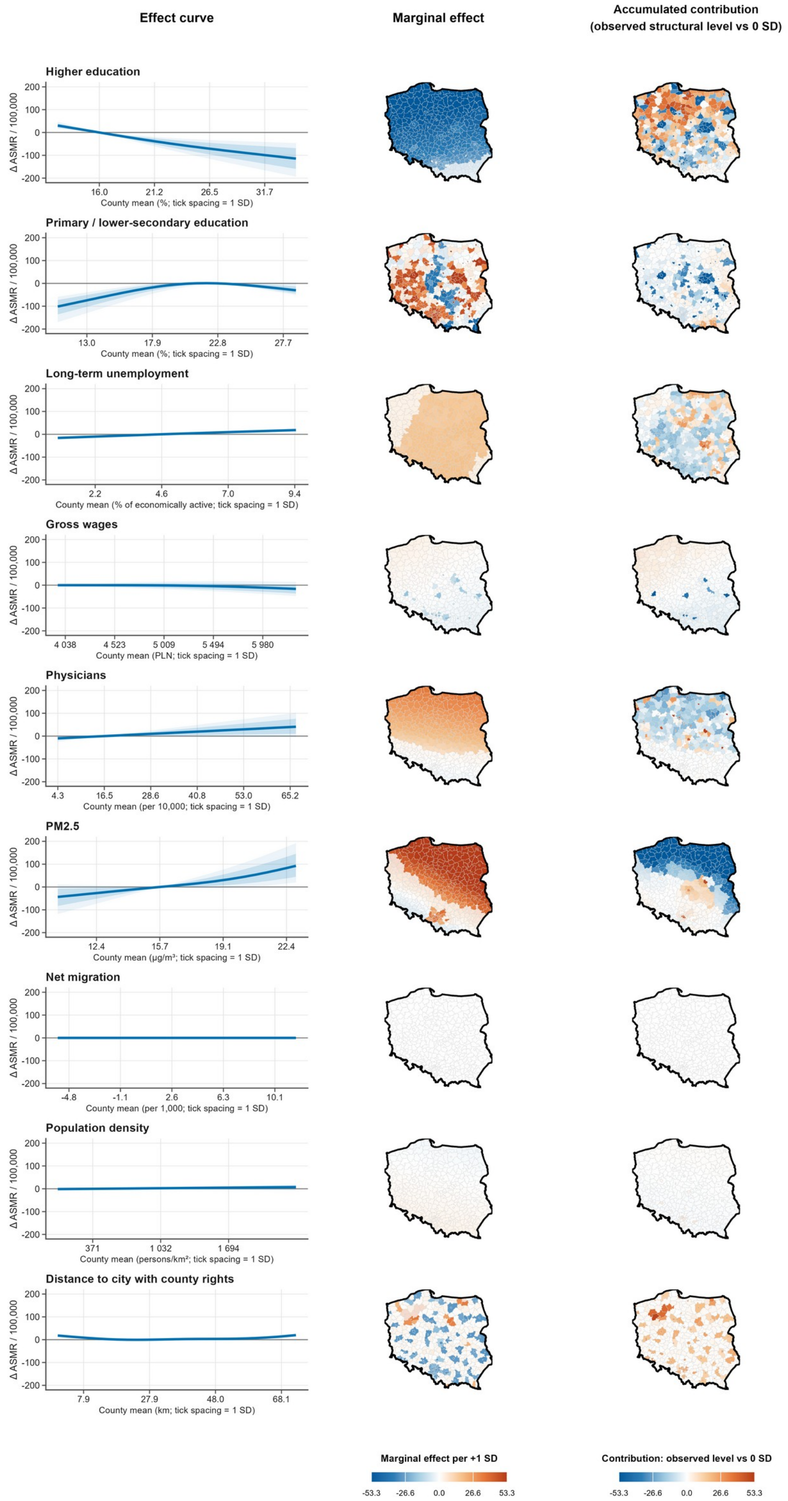


**Supplementary Figure S4.** Full Model 3 between-county effect grid, 2010–2025. Map lines delineate study areas and do not necessarily depict accepted national boundaries.

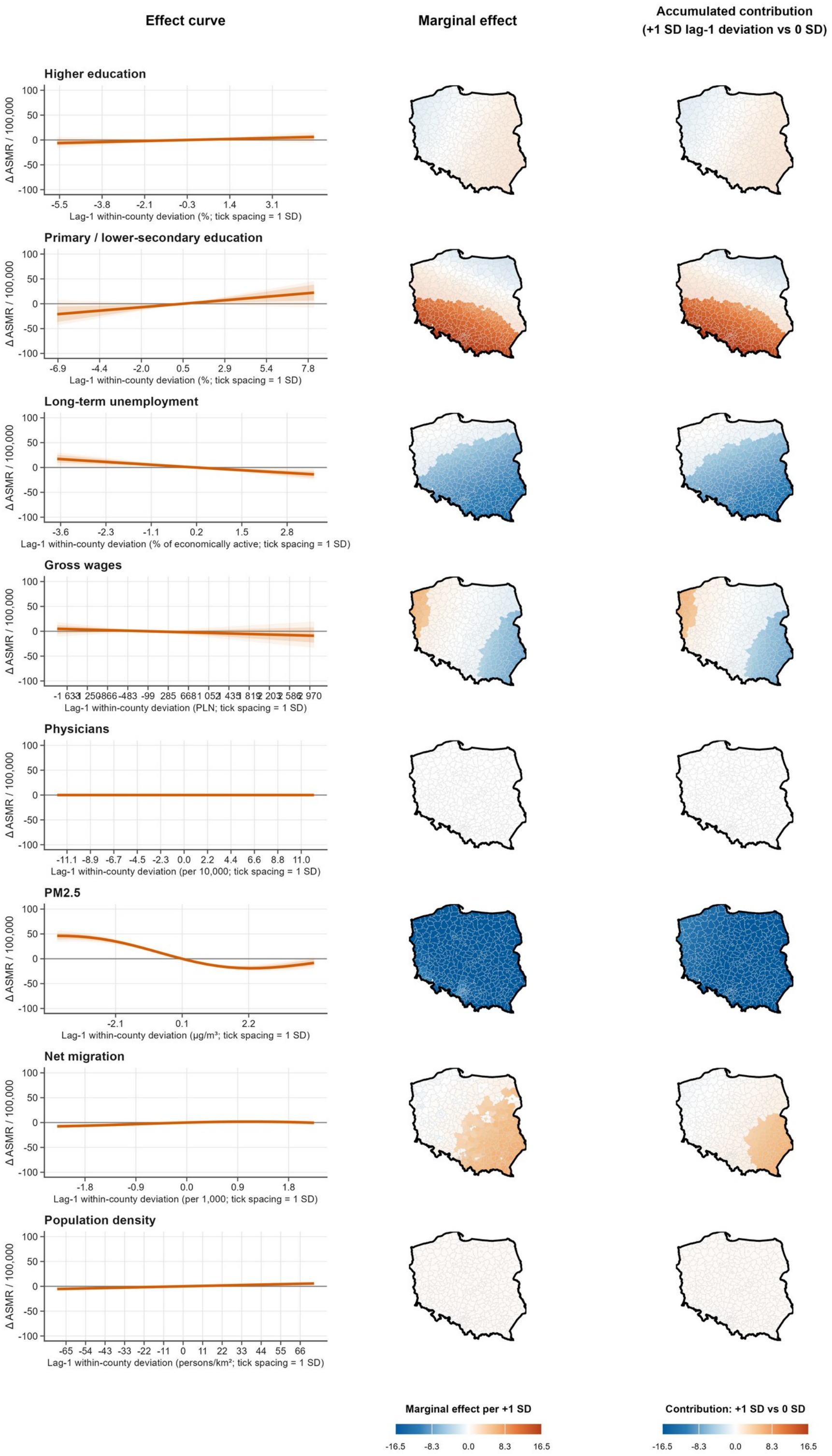


**Supplementary Figure S5.** Full Model 3 within-county lag-1 effect grid, 2010–2025. Map lines delineate study areas and do not necessarily depict accepted national boundaries.

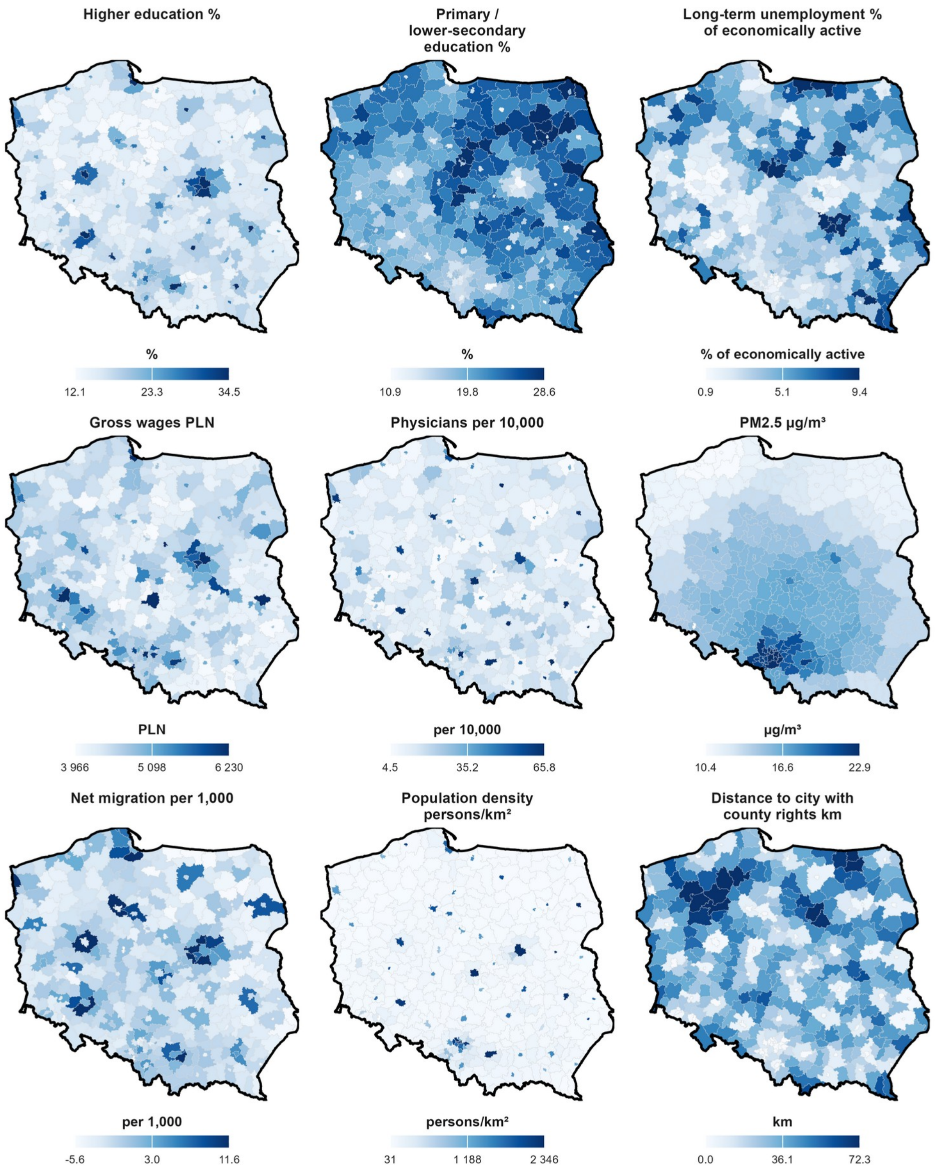


**Supplementary Figure S6.** Mean raw determinant values by county, 2010–2025. Map lines delineate study areas and do not necessarily depict accepted national boundaries.

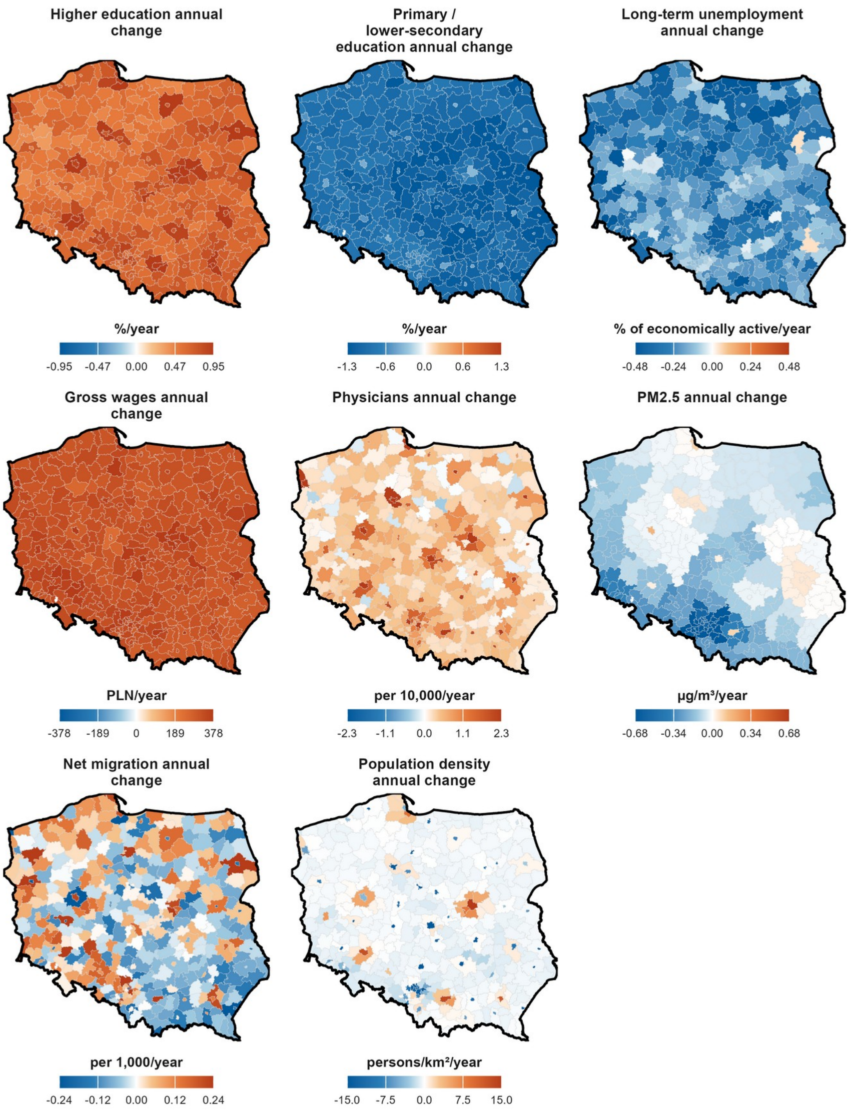


**Supplementary Figure S7.** County-specific annual changes in raw time-varying determinants, 2010–2025. Map lines delineate study areas and do not necessarily depict accepted national boundaries.

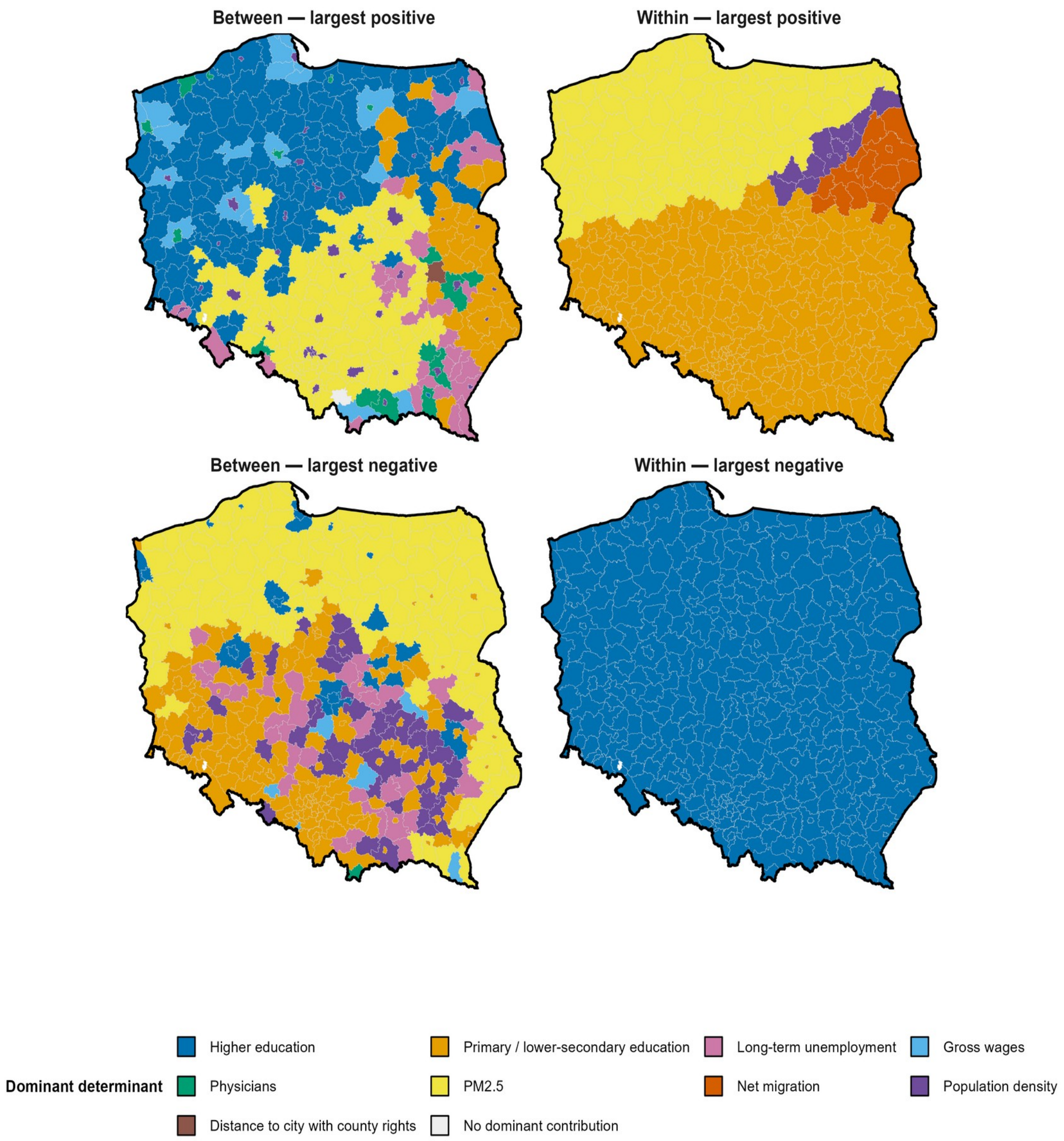


**Supplementary Figure S8.** Dominant accumulated determinant contribution by county and sign, Model 2 (2010–2022). Model 1 is shown in main-text Figure 5. Dominance identifies the largest reference-based accumulated contribution within each county and does not rank determinants by national effect magnitude. Within-county accumulated contributions use the +1 SD lag-1 deviation versus 0-SD reference contrast. Map lines delineate study areas and do not necessarily depict accepted national boundaries.

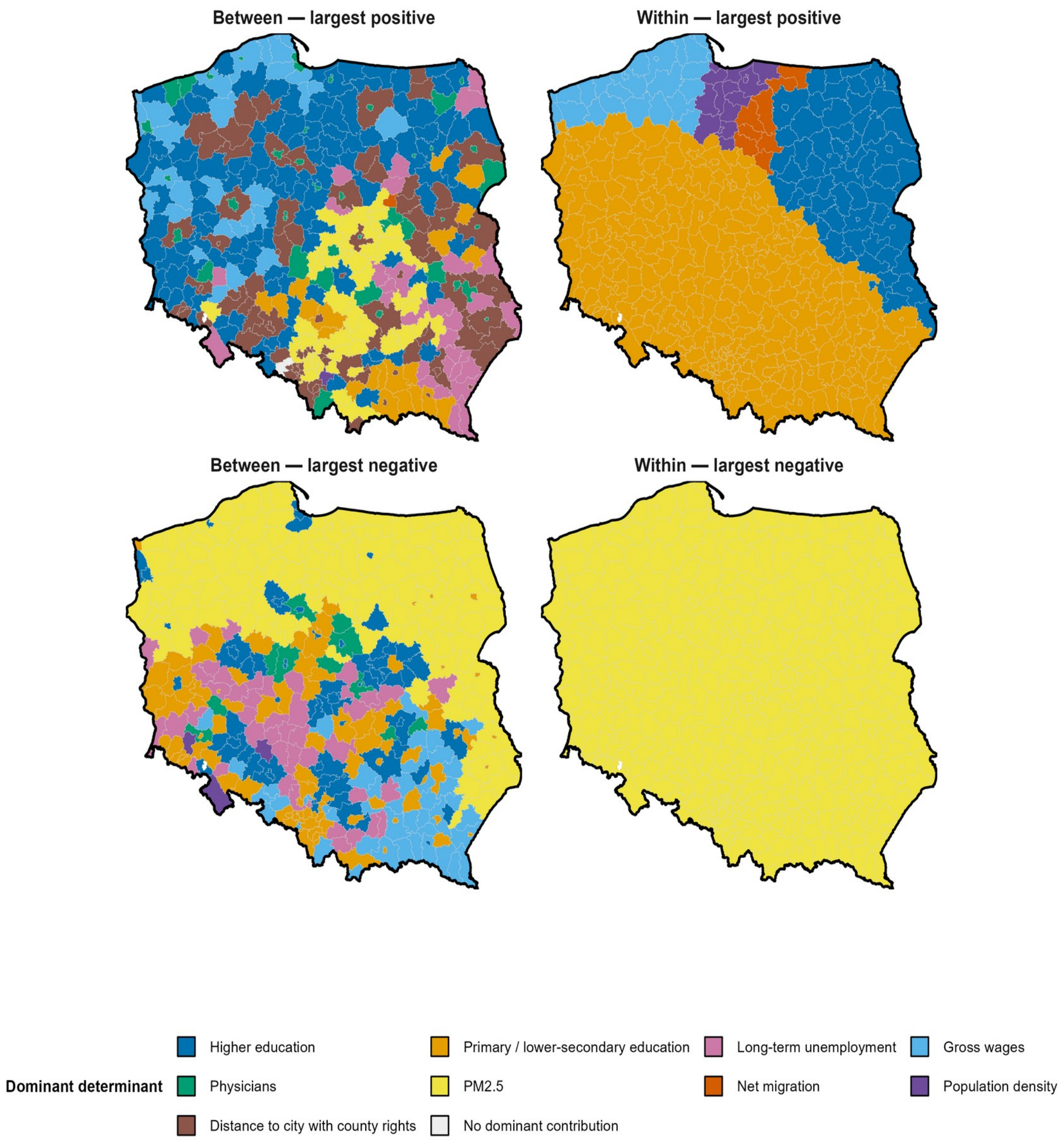


**Supplementary Figure S9.** Dominant accumulated determinant contribution by county and sign, Model 3 (2010–2025). Dominance identifies the largest reference-based accumulated contribution within each county and does not rank determinants by national effect magnitude. Within-county accumulated contributions use the +1 SD lag-1 deviation versus 0-SD reference contrast. Map lines delineate study areas and do not necessarily depict accepted national boundaries.

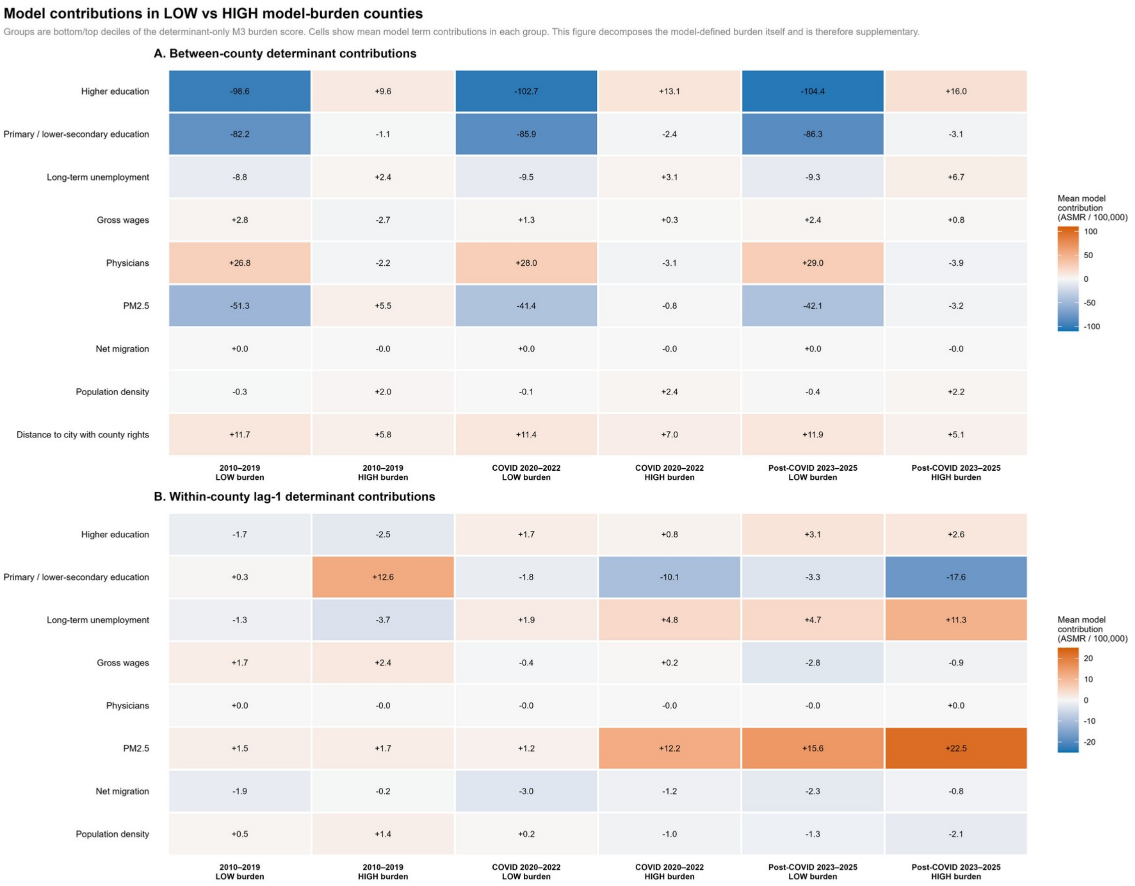


**Supplementary Figure S10.** Sensitivity profile using LOW versus HIGH model-defined determinant-burden counties rather than observed-mortality groups. This figure decomposes the model score itself and is secondary to the outcome-defined comparison in Supplementary Tables S7–S8.

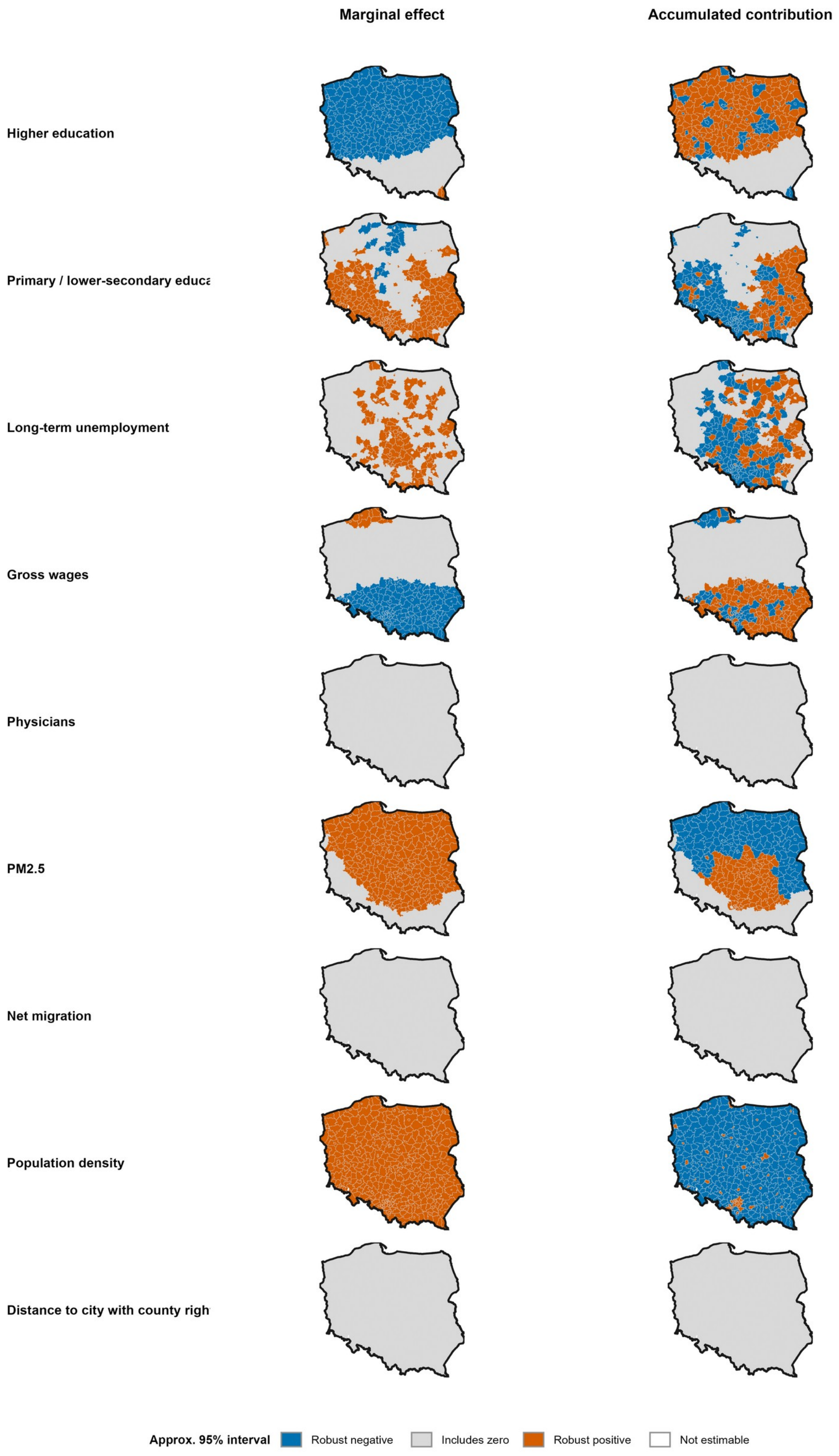


**Supplementary Figure S11.** Local coefficient-covariance uncertainty for Model 1 between-county marginal effects and accumulated contributions. Blue = robust negative; grey = interval includes zero; orange = robust positive; white = not estimable. Robust sign means the approximate pointwise 95% interval excludes zero; intervals are not multiplicity-adjusted and are not causal significance tests. Map lines delineate study areas and do not necessarily depict accepted national boundaries.

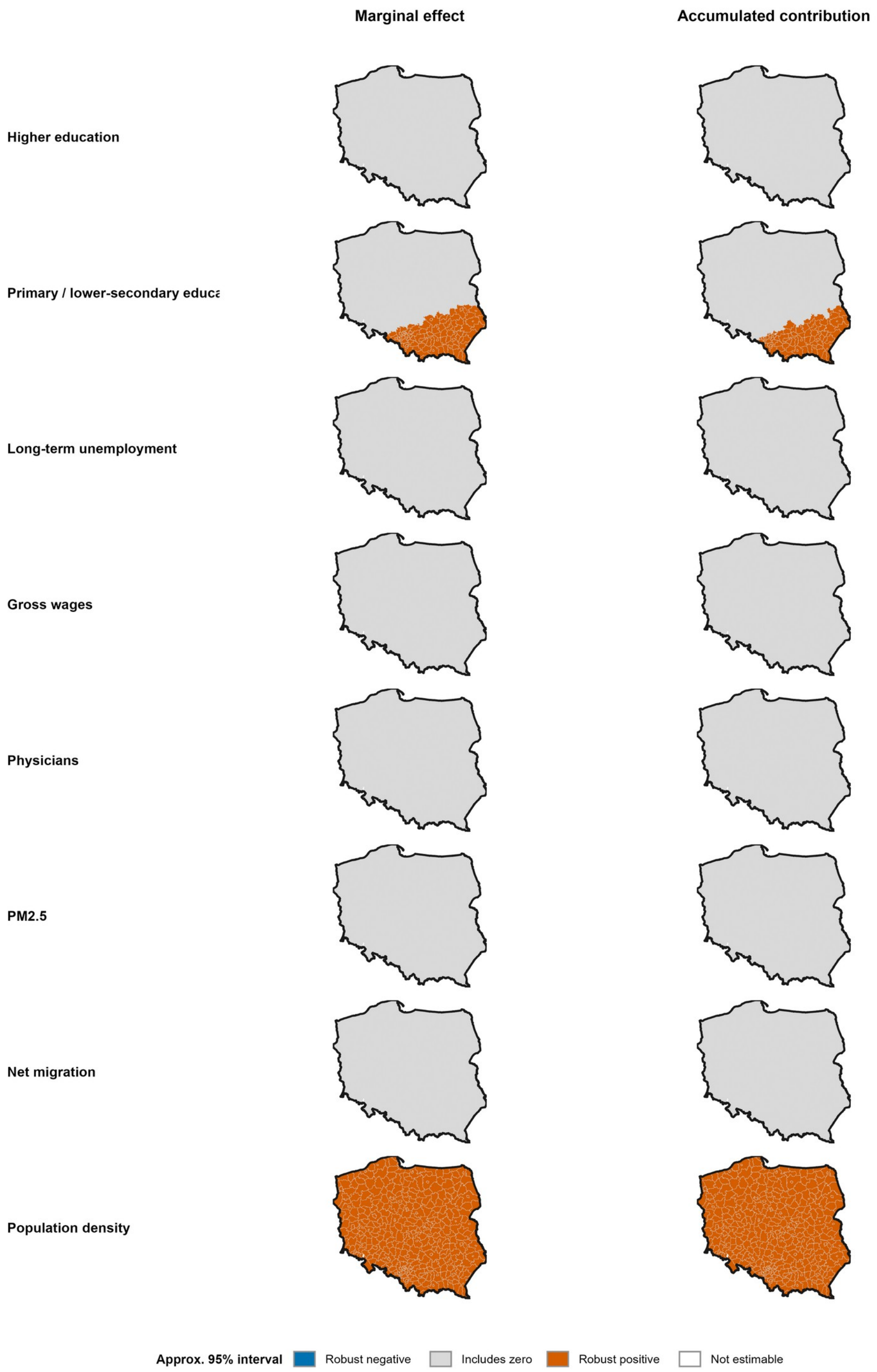


**Supplementary Figure S12.** Local coefficient-covariance uncertainty for Model 1 within-county lag-1 marginal effects and accumulated contributions. Blue = robust negative; grey = interval includes zero; orange = robust positive; white = not estimable. Most within-county local surfaces are weakly identified in Model 1, which is distinct from their global/APS or perturbation behaviour. Map lines delineate study areas and do not necessarily depict accepted national boundaries.

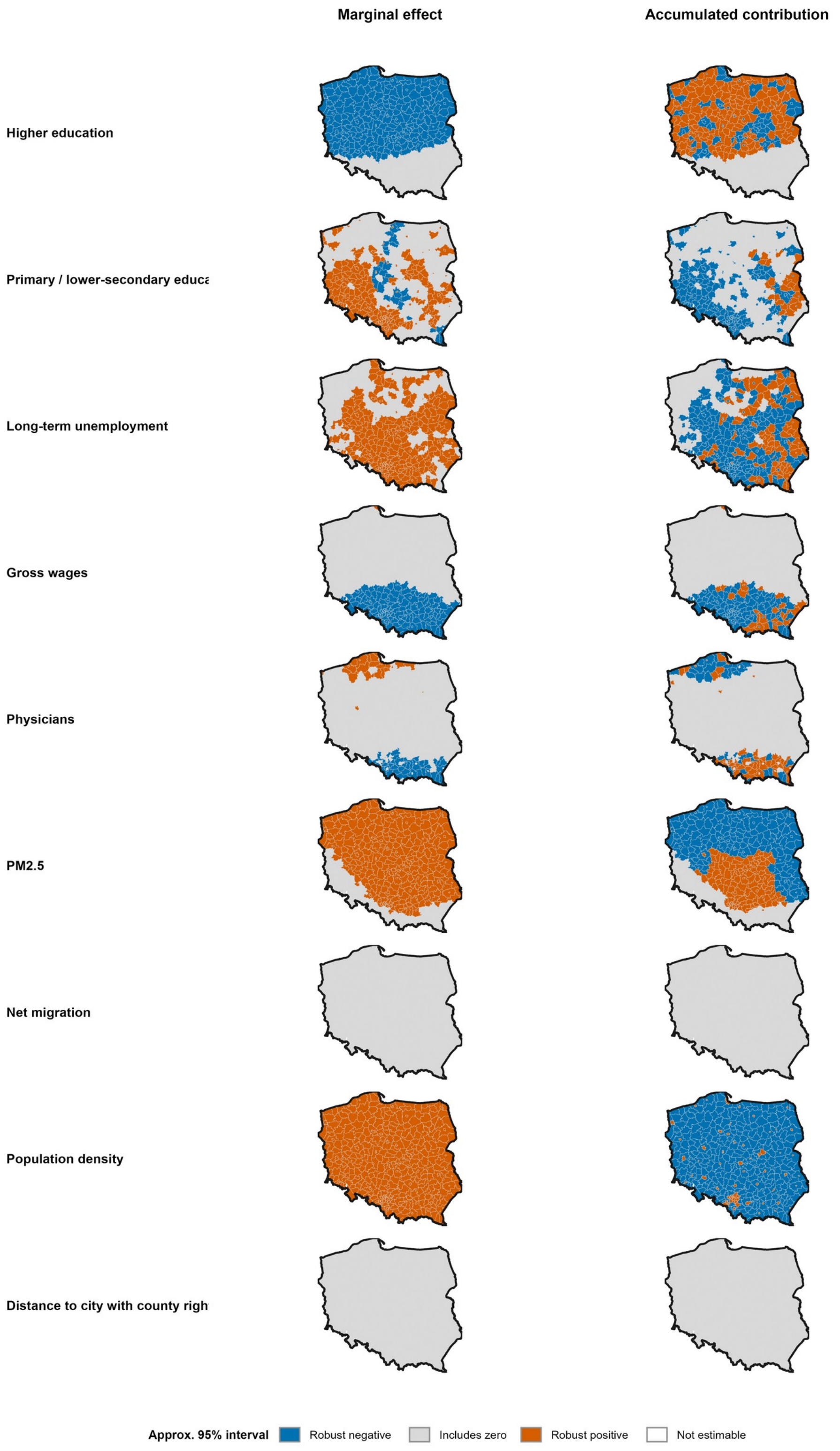


**Supplementary Figure S13.** Local coefficient-covariance uncertainty for Model 2 between-county marginal effects and accumulated contributions. Blue = robust negative; grey = interval includes zero; orange = robust positive; white = not estimable. Robust sign means the approximate pointwise 95% coefficient-covariance interval excludes zero; intervals are descriptive, not multiplicity-adjusted, and are not causal significance tests. Map lines delineate study areas and do not necessarily depict accepted national boundaries.

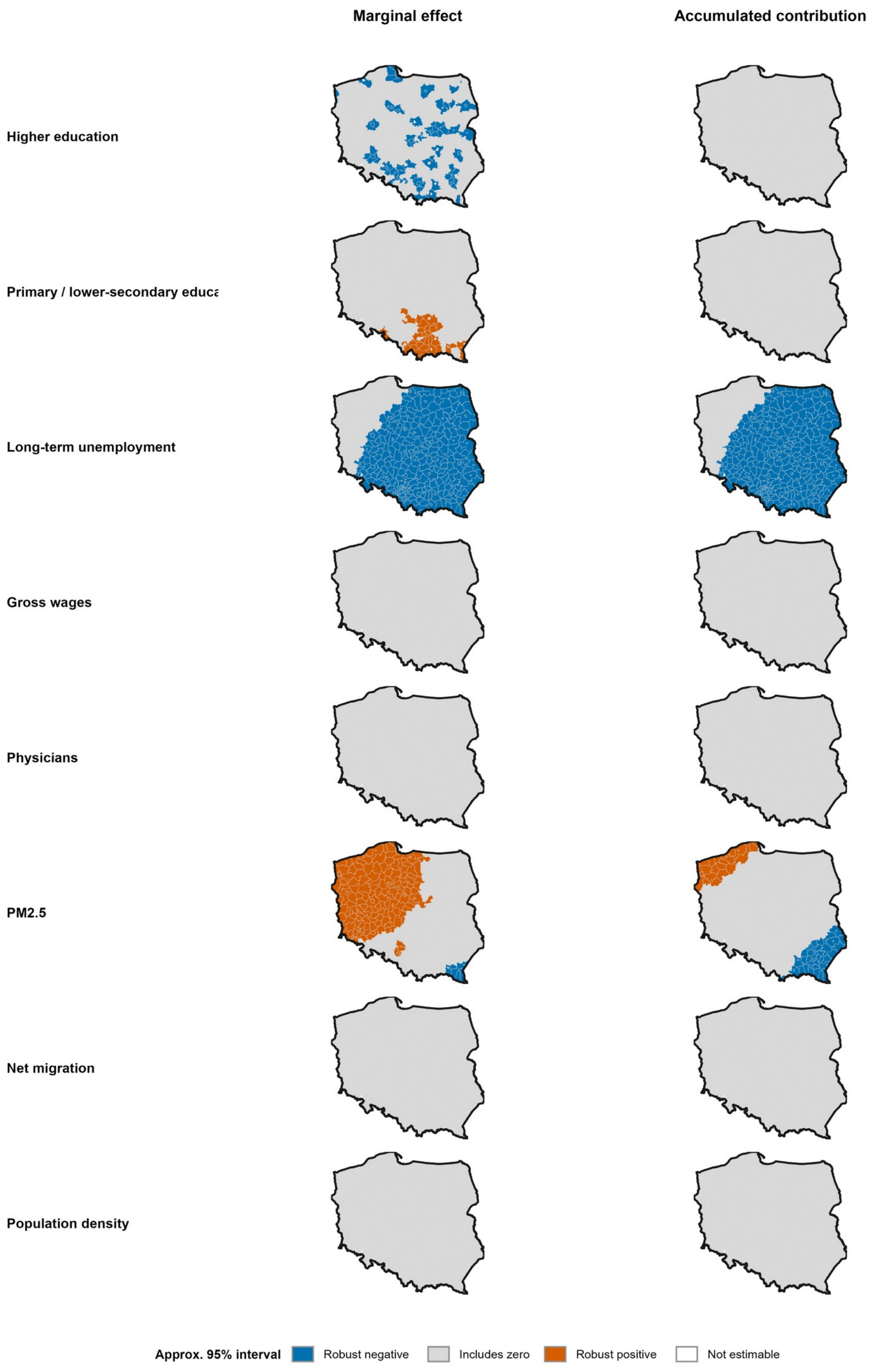


**Supplementary Figure S14.** Local coefficient-covariance uncertainty for Model 2 within-county lag-1 marginal effects and accumulated contributions. Classification uses approximate pointwise 95% coefficient-covariance intervals and is descriptive rather than multiplicity-adjusted. Map lines delineate study areas and do not necessarily depict accepted national boundaries.

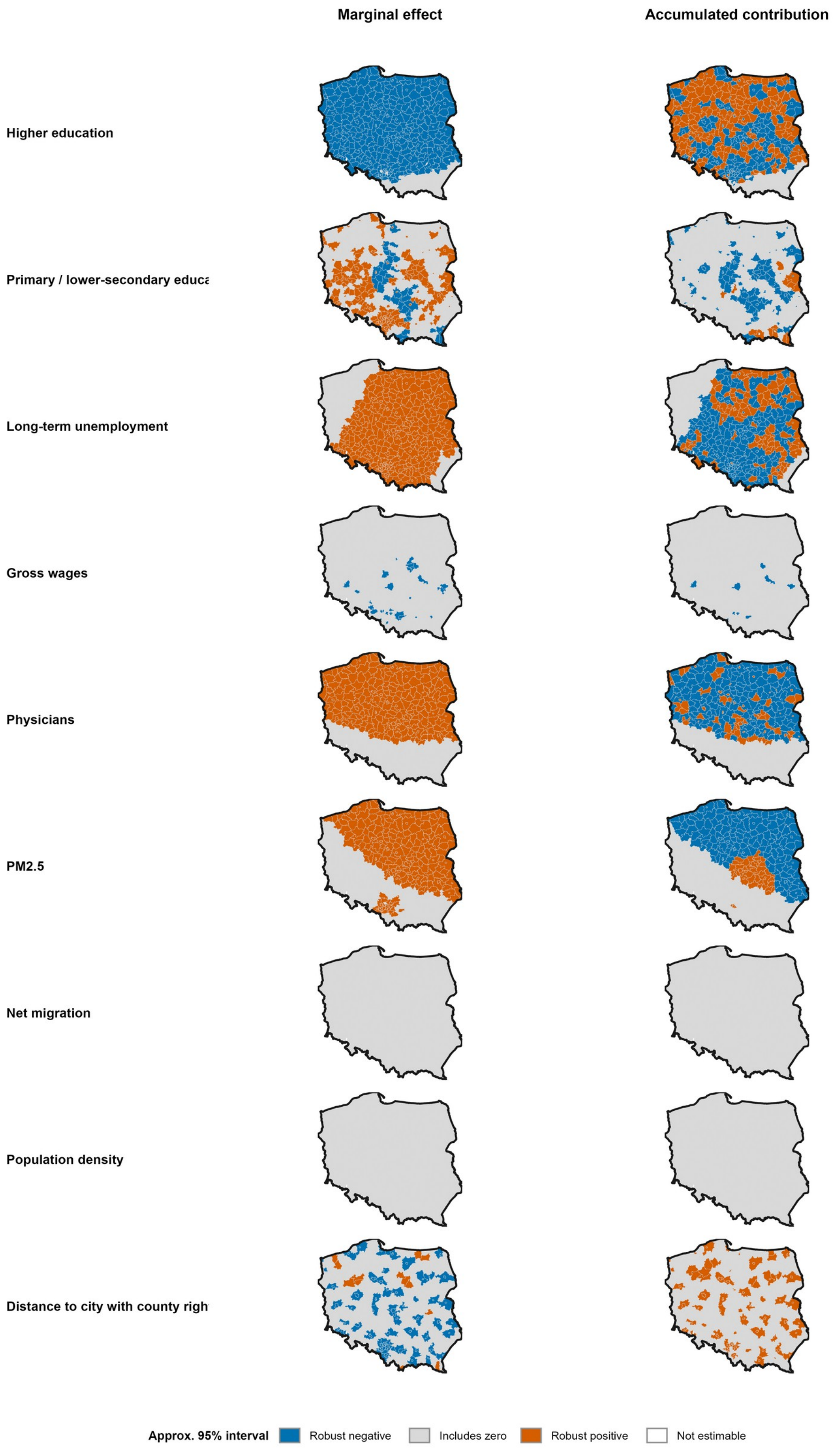


**Supplementary Figure S15.** Local coefficient-covariance uncertainty for Model 3 between-county marginal effects and accumulated contributions. Classification uses approximate pointwise 95% coefficient-covariance intervals and is descriptive rather than multiplicity-adjusted. Map lines delineate study areas and do not necessarily depict accepted national boundaries.

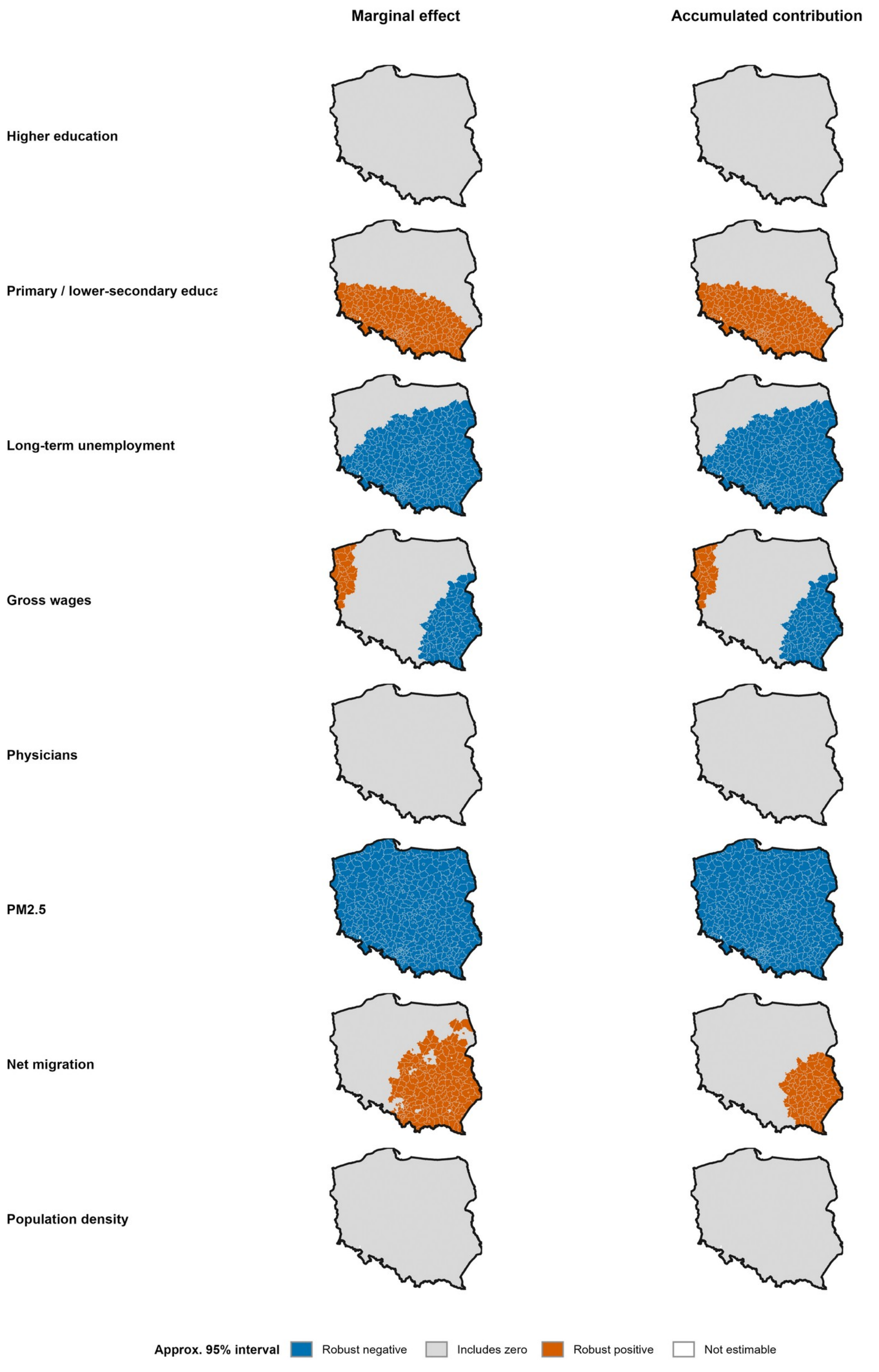


**Supplementary Figure S16.** Local coefficient-covariance uncertainty for Model 3 within-county lag-1 marginal effects and accumulated contributions. Classification uses approximate pointwise 95% coefficient-covariance intervals and is descriptive rather than multiplicity-adjusted. Map lines delineate study areas and do not necessarily depict accepted national boundaries.

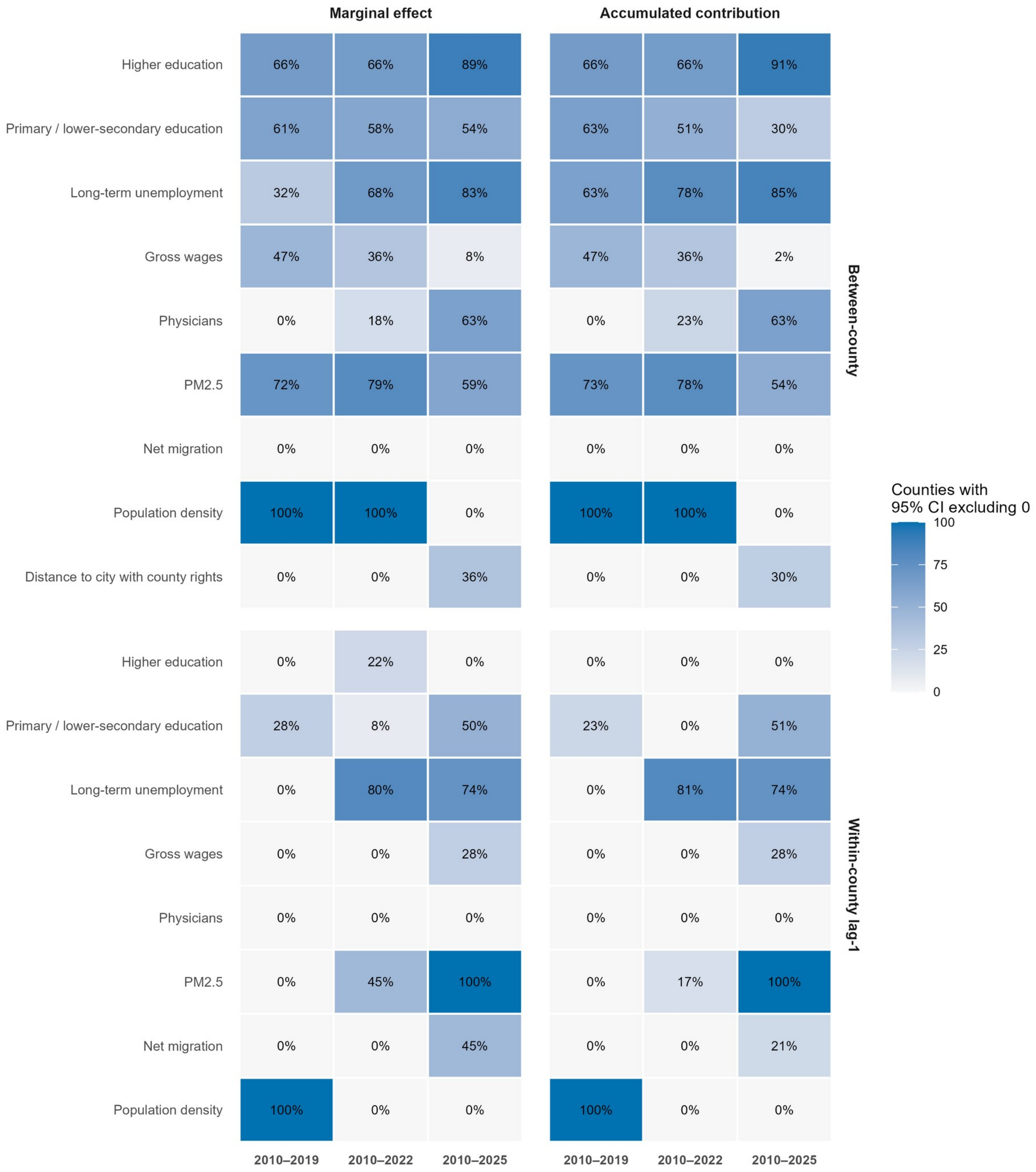


**Supplementary Figure S17.** Local uncertainty coverage across Models 1–3. Cells report the percentage of counties whose approximate pointwise 95% coefficient-covariance interval excludes zero for each determinant, component and map contrast. This is an uncertainty summary, not a multiplicity-adjusted significance analysis.

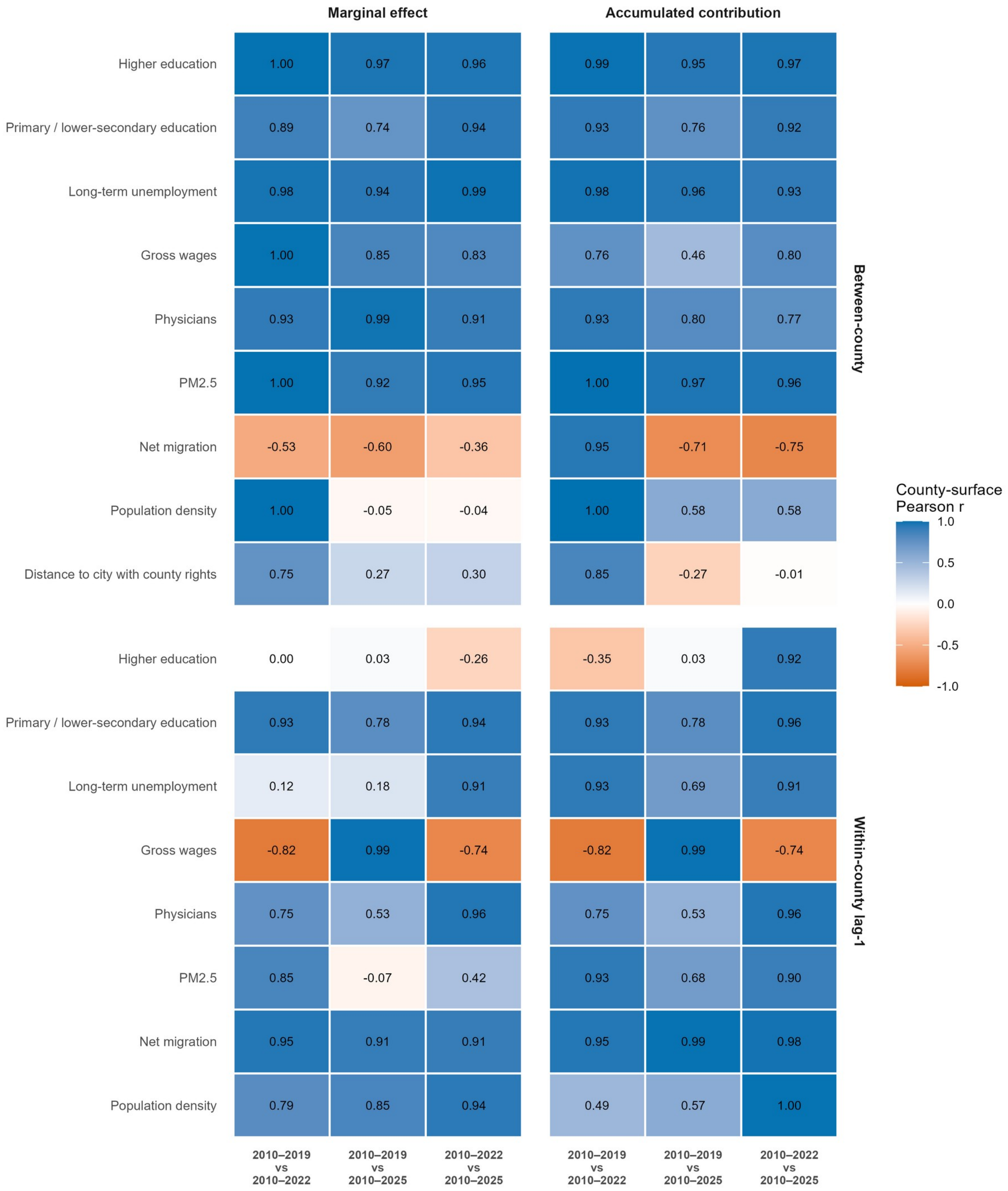


**Supplementary Figure S18.** Cross-period stability of county-specific SVC effect surfaces. Correlations compare overlapping nested model windows and quantify sensitivity to extending the analytical period; they should not be interpreted as independent replication across periods.
